# Associations between SARS-CoV-2 infection and incidence of new chronic condition diagnoses: a systematic review

**DOI:** 10.1101/2023.02.21.23286181

**Authors:** Lindsay A. Gaudet, Jennifer Pillay, Sabrina Saba, Dianne Zakaria, Nicholas Cheta, Hélène Gardiner, Larry Shaver, Jacqueline Middleton, Maria Tan, Ben Vandermeer, Lisa Hartling

**Affiliations:** Alberta Research Center for Health Evidence, University of Alberta, Edmonton, AB, Canada; Public Health Agency of Canada, Ottawa, ON, Canada; Epidemiology Coordinating and Research Centre, University of Alberta, Edmonton, AB, Canada

**Keywords:** COVID-19, SARS-CoV-2, incidence, chronic conditions, systematic review, meta-analysis

## Abstract

Because of the large number of infected individuals, an estimate of the future burdens of the long-term consequences of SARS-CoV-2 infection is needed. This systematic review examined associations between SARS-CoV-2 infection and incidence of categories of and selected chronic conditions, by age and severity of infection (inpatient vs. outpatient/mixed care). MEDLINE and EMBASE were searched (Jan 1, 2020 to Oct 4, 2022) and reference lists scanned. We included observational studies from high-income OECD countries with a control group adjusting for sex and comorbidities. Identified records underwent a two-stage screening process. Two reviewers screened 50% of titles/abstracts, after which DistillerAI acted as second reviewer. Two reviewers then screened the full texts of stage one selections. One reviewer extracted data and assessed risk of bias; results were verified by another. Random-effects meta-analysis estimated pooled hazard ratios (HR). GRADE assessed certainty of the evidence. Twenty-five studies were included. Among the outpatient/mixed SARS-CoV-2 care group, there is high certainty of a small-to-moderate increase (i.e., HR 1.26 to 1.99) among adults ≥65 years of any cardiovascular condition, and of little-to-no difference (i.e., HR 0.75 to 1.25) in anxiety disorders for individuals <18, 18-64, and ≥65 years old. Among 18-64 and ≥65 year-olds receiving outpatient/mixed care there are probably (moderate certainty) large increases (i.e., HR ≥2.0) in encephalopathy, interstitial lung disease, and respiratory failure. After SARS-CoV-2 infection, there is probably an increased risk of diagnoses for some chronic conditions; whether the magnitude of risk will remain stable into the future is uncertain.

## Introduction

In addition to disrupting the global economy, [1] SARS-CoV-2 has infected millions of people worldwide and more than 4.5 million Canadians. [2] Potential long-term consequences of SARS-CoV-2 infection were raised in the first year of the pandemic. [3] Combined with the large number of infected individuals, it is necessary to derive some estimate of the future burdens of the long-term consequences of SARS-CoV-2 infection so that health policy and other decision makers can make informed decisions and healthcare systems can prepare for a potential increase in need for care and resources.

Many reviews in the literature have examined post-COVID-19 condition (previously called Long COVID), [4–6] and many reviews reporting on other long-term sequalae, such as the development of chronic conditions after SARS-CoV-2 infection, have been limited to a single condition or cluster of conditions [7–9] and/or did not require included studies to have a control group in order to quantify attributable risk. [10–12] In order to understand how SARS-CoV-2 may change the future burden of health outcomes on healthcare resources in the future, it is important to assess whether there is actually an association between SARS-CoV-2 infection and increased risk of long-term sequelae.

Therefore, we set out to conduct a systematic review to answer the question: What are the associations between SARS-CoV-2 infection and the incidence of new diagnoses or exacerbations of chronic conditions in groups based on age and severity of infection?

## Methods

This review followed an *a priori* protocol developed in consultation with disease leads (NC, DZ, LS, HG, JM, and others) at the Public Health Agency of Canada. The protocol was prospectively registered and is available on PROSPERO (CRD42022364883). This review has been reported according to the Preferred Reporting Items for Systematic Reviews and Meta-analyses 2020 reporting guideline (Appendix 1 in the supplement). [13] Data are available from the authors on reasonable request.

The authors have no competing interests to declare.

### Study Eligibility

We included prospective or retrospective observational studies carried out in high-income Organisation for Economic Co-operation and Development (OECD) member countries [14] and comparing individuals with suspected or confirmed SARS-CoV-2 infection (exposed) to those without (controls). Pre-prints and other reports not peer-reviewed were eligible. Conference abstracts frequently present preliminary results and rarely report sufficient methods to adequately assess quality and were therefore excluded. We limited inclusion to records published in English or French, as these are the official languages of Canada and limits on non-English language studies has not been shown to bias systematic review conclusions. [15] Table S1 in the Supplement outlines our eligibility criteria in greater detail.

**Table 1.** Study characteristics of included studies for a systematic review of new diagnoses of chronic conditions after SARS-CoV-2 infection.

| Study<br>Country<br>Data source | Index time<br>for SARS-CoV-2<br>infections | Study design | Mean/ median<br>follow-up, range<br>If not reported,<br>maximum<br>range of FU? | Care<br>type/setting<br>for SARS-CoV-2<br>infected<br>cases | Age range<br>(years)<br>% Female | % Hospitalized<br>% in ICU | Comparator timing<br>Test status of<br>comparator group | Outcomes |
| --- | --- | --- | --- | --- | --- | --- | --- | --- |
| Abel 2021 [32]<br>United Kingdom<br>Clinical Practice<br>Research<br>Datalink Aurum | Feb 2020 to<br>Dec 2020 | Retrospective<br>cohort | Median (IQR) –<br>6.3 (4.0-9.3) weeks | Outpatient | 16 to 80+<br>50% | NA | Concurrent<br>No<br>confirmed/suspected<br>SARS-CoV-2<br>infection | Mental disorders (anxiety disorders,<br>depression, psychosis) |
| Ayoubkhani 2021<br>[33]<br>United Kingdom<br>Hospital Episode<br>Statistics Admitted<br>Patient Care and<br>General Practice<br>Extraction Service<br>Data for Pandemic<br>Planning and<br>Research | Jan 2020 to<br>Aug 2020 | Retrospective<br>cohort | Cov: Mean (SD) –<br>140 (50) days<br>Con: Mean (SD) –<br>153 (33) days | Inpatient | 0 to 70 +<br>45% | 100<br>NR | Concurrent<br>No<br>confirmed/suspected<br>SARS-CoV-2<br>infection | Cardiovascular disease (composite of<br>arrhythmia, heart failure, myocardial<br>infarction, and stroke); Chronic<br>kidney disease (dialysis and kidney<br>transplant); Diabetes (type 1 & type<br>2); Respiratory disorders |
| Bohlken 2022 [34]<br>Germany<br>IQVIA Disease<br>Analyzer database | Mar 2020 to<br>Sept 2021 | Retrospective<br>cohort | Cov: Mean – 158<br>days<br>Con: Mean – 165<br>days | Outpatient | 18 to 70+<br>53.3% | NA | Concurrent<br>>90% had test<br>negative | Neurological conditions (mild<br>cognitive disorder) |

| Study<br>Country<br>Data source | Index time<br>for SARS-CoV-2<br>infections | Study design | Mean/ median<br>follow-up, range<br>If not reported,<br>maximum<br>range of FU? | Care<br>type/setting<br>for SARS-CoV-2<br>infected<br>cases | Age range<br>(years)<br>% Female | %<br>Hospitalized<br>% in ICU | Comparator timing<br>Test status of<br>comparator group | Outcomes |
| --- | --- | --- | --- | --- | --- | --- | --- | --- |
| Chevinsky 2021<br>[35]<br>United States<br>Premier Healthcare<br>Database Special<br>COVID-19 Release | Mar 2020 to<br>Jun 2020 | Prospective<br>cohort | Range 1 to 4 mos | Mixed | <18 (adult data<br>not stratified by<br>age) | 11.4<br>NR | Concurrent<br>No<br>confirmed/suspected<br>SARS-CoV-2<br>infection | 31 different conditions across 7<br>categories, but only reports<br>“Children with COVID-19 were not<br>more likely to experience new<br>diagnoses than children without<br>COVID-19”, with no effect size or<br>variance reported and was thus<br>unable to be included in the meta-analysis. Attempt to contact authors<br>to obtain the data was not successful. |
| Cohen 2022 [36]<br>United States<br>UnitedHealth<br>Database | Jan 2020 to<br>Dec 2020 | Retrospective<br>cohort | Median (IQR) – 78<br>(30 – 175) days | Mixed | 65+<br>58% | 27<br>6.4 | Concurrent<br>No<br>confirmed/suspected<br>SARS-CoV-2<br>infection | Cardiovascular disease (acute<br>coronary disease, cardiogenic shock,<br>cardiac arrhythmia, cardiomyopathy,<br>congestive heart failure, coronary<br>disease, hypertension, myocardial<br>infarction, tachycardia); Chronic<br>kidney disease; Diabetes (type 2);<br>Mental disorders (mental health<br>diagnosis, psychosis); Neurological<br>conditions (dementia,<br>encephalopathy, Guillain-Barre<br>syndrome, migraine, peripheral<br>neuropathy, seizure); Respiratory<br>disorders (chronic respiratory failure,<br>interstitial lung disease); Stroke |
| Daugherty 2021<br>[37]<br>United States<br>UnitedHealth<br>Database | Jan 2019 to<br>Oct 2020 | Retrospective<br>cohort | Median (IQR) –<br>87 (45-124) days | Mixed | 18 to 65<br>52.5% | 8.2<br>1.1 | Concurrent<br>No<br>confirmed/suspected<br>SARS-CoV-2<br>infection | Cardiovascular disease (acute<br>coronary disease, arrhythmia,<br>cardiogenic shock, cardiomyopathy,<br>congestive heart failure, coronary<br>disease, hypertension, myocardial<br>infarction, tachycardia); Chronic<br>kidney disease; Diabetes (type 2);<br>Mental disorders (mental health<br>diagnosis, psychosis); Neurological<br>conditions (Alzheimer’s disease,<br>dementia, encephalopathy, Guillain-Barre syndrome, migraine, peripheral<br>neuropathy, seizure); Respiratory<br>disorders (chronic respiratory failure,<br>interstitial lung disease); Stroke |
| Donnachie 2022<br>[38]<br>Germany<br>Bavarian COVID-19<br>Cohort | Jan 2020 to Jun<br>2021 | Retrospective<br>cohort | NR<br>Followed up for 2<br>years | Outpatient | 0 to 60+<br>54% | NA | Concurrent<br>>90% had test<br>negative | Mental disorders (anxiety disorders,<br>mood disorders); Neurological<br>conditions (mild cognitive<br>impairment) |
| Jacob 2022 [39]<br>Germany<br>IQVIA Disease<br>Analyzer database | Mar 2020 to<br>May 2021 | Retrospective<br>cohort | NR<br>Maximum of 14<br>mos | Outpatient | 18 to 70+<br>52.3% | NA | Concurrent<br>No<br>confirmed/suspected<br>SARS-CoV-2<br>infection | Mental disorders (anxiety disorders,<br>depression) |
| Kompaniyets 2022<br>[40]<br>United States<br>HealthVerity | Mar 2020 to<br>Jan 2022 | Retrospective<br>cohort | NR<br>Minimum of 60<br>days to maximum<br>of 365 days | Mixed | 2 to 17<br>50% | NR | Concurrent<br>No<br>confirmed/suspected<br>SARS-CoV-2<br>infection | Cardiovascular disease (cardiac<br>dysrhythmias); Chronic kidney<br>disease; Diabetes; Mental disorders<br>(anxiety disorders, mood disorder);<br>Musculoskeletal disorders;<br>Neurological conditions (nervous<br>system disorder); Respiratory<br>disorders (asthma); Stroke |

| **Study**<br>**Country**<br>**Data source** | **Index time**<br>**for SARS-**<br>**CoV-2**<br>**infections** | **Study design** | **Mean/ median**<br>**follow-up, range**<br>**If not reported,**<br>**maximum**<br>**range of FU?** | **Care**<br>**type/setting**<br>**for SARS-**<br>**CoV-2**<br>**infected**<br>**cases** | **Age range**<br>**(years)**<br>**% Female** | **% Hospitalized**<br>**% in ICU** | **Comparator timing**<br>**Test status of**<br>**comparator group** | **Outcomes** |
| --- | --- | --- | --- | --- | --- | --- | --- | --- |
| Park 2021 [41]<br>Korea<br>National Health<br>Insurance Service<br>Database | Jan 2020 to<br>Dec 2020 | Retrospective<br>cohort | NR<br>Minimum of 0<br>days to maximum<br>of 12 mos | Mixed | 20 to 60+<br>54.3% | NR | Concurrent<br>No<br>confirmed/suspected<br>SARS-CoV-2<br>infection | Mental disorders (mental illness) |
| Pietropaolo 2022 [42]<br>United States<br>TriNetX COVID-19<br>Research Network | Jan 2020 to Jun<br>2021 | Retrospective<br>cohort | NR<br>Minimum of 1 day<br>to maximum of 18<br>mos | Mixed | 0 to 30<br>45% | NR | Concurrent<br>No<br>confirmed/suspected<br>SARS-CoV-2<br>infection | Diabetes (type 1 & type 2) |
| Qureshi 2022 [43]<br>United States<br>Cerner Real-World<br>Data | Until July 2021 | Retrospective<br>cohort | Median (IQR) –<br>182 (113 – 277)<br>days | Inpatient | 0 to 70+<br>39% | 100<br>NR | Concurrent<br>>90% had test<br>negative | Neurological conditions (dementia) |
| Rao 2022 [44]<br>US<br>Electronic health<br>record data from<br>PEDSnet<br>institutions | Mar 2020 to<br>Oct 2021 | Retrospective<br>cohort | Cov: Mean (SD) –<br>4.6 (0.7) Weeks<br>Con: Mean (SD) –<br>4.7 (0.7) weeks | Mixed | 0 to 21<br>47.2% | 6<br>2.2 | Concurrent<br>>90% had test<br>negative | Mental disorders (mental health<br>treatment); Neurological conditions<br>(communication/motor disorders) |
| Rezel-Potts 2022 [45]<br>United Kingdom<br>Clinical Practice<br>Research<br>Datalink Aurum | Feb 2021 to<br>Jan 2022 | Retrospective<br>cohort | Median – 12 mos | Mixed | ---<br>56% | NR | Concurrent<br>No<br>confirmed/suspected<br>SARS-CoV-2<br>infection | Cardiovascular disease (atrial<br>arrhythmias, heart failure,<br>myocardial infarction and ischemic<br>heart disease); Diabetes (type 1 &<br>type 2); Stroke |

| Study<br>Country<br>Data source | Index time<br>for SARS-CoV-2<br>infections | Study design | Mean/ median<br>follow-up, range<br>If not reported,<br>maximum<br>range of FU? | Care<br>type/setting<br>for SARS-CoV-2<br>infected<br>cases | Age range<br>(years)<br>% Female | %<br>Hospitalize<br>d<br>% in ICU | Comparator timing<br>Test status of<br>comparator group | Outcomes |
| --- | --- | --- | --- | --- | --- | --- | --- | --- |
| Roessler 2022 [46]<br>Germany<br>Data from 6<br>German statutory<br>health insurance<br>organizations: AOK<br>Bayern —Die<br>Gesundheitskasse,<br>AOK PLUS,<br>BARMER, BKKen,<br>DAK Gesundheit,<br>and Techniker<br>Krankenkasse | By Jun 2020 | Retrospective<br>cohort | Cov: Mean (SD) –<br>236 (44) days<br>Con: Mean (SD) –<br>254 (36) days | Mixed | 0 to 18<br>48.1% | 1<br>0.4 | Concurrent<br>No<br>confirmed/suspected<br>SARS-CoV-2<br>infection | Cardiovascular disease (cardiac<br>arrhythmias, heart failure, heart<br>murmurs, myocardial infarction,<br>other cardiac arrhythmias); Mental<br>disorders (adjustment disorder,<br>anxiety disorders, depressive<br>disorders, emotional and behavioural<br>disorders, obsessive-compulsive<br>disorder); Neurological conditions<br>(chronic fatigue syndrome,<br>developmental delay, dyslexia, facial<br>nerve paralysis, headache, movement<br>disorders, other coordination<br>disorders/ataxia, seizures, speech and<br>language disorders); Stroke |
| Tartof 2022 [16]<br>United States<br>Vaccine Safety<br>Datalink | Mar 2019 to<br>Mar 2021 | Retrospective<br>cohort | NR<br>Maximum of 6<br>mos | Inpatient | 0 to 85+<br>53.7% | 100<br>NR | Concurrent<br>>90% had test<br>negative | Diabetes; Mental disorder (anxiety<br>disorders, psychosis); Stroke<br>Only <18y data eligible for meta-analysis |
| Taquet 2022 [47]<br>United States<br>TriNetX COVID-19<br>Research Network | Jan 2020 to<br>Mar 2022 | Retrospective<br>cohort | Cov: Mean (SD) –<br>213 (204) days<br>Con: Mean (SD) –<br>223 (203) days | Mixed | 0 to 65+<br>57.8% | NR | Concurrent<br>>90% had test<br>negative | Mental disorders (anxiety disorders,<br>mood disorders, psychotic disorder);<br>Neurological conditions (cognitive<br>deficit, dementia, Guillain-Barre<br>syndrome, myoneural<br>junction/muscle disease, nerve/nerve<br>root/plexus disorder, Parkinsonism,<br>seizure); Stroke |
| Taquet 2021 [48]<br>United States<br>TriNetX COVID-19<br>Research Network | Jan 2020 to<br>Apr 2022 | Retrospective<br>cohort | NR<br>14 days to 90 days | Mixed | 65+ | NR | Concurrent<br>>90% had test<br>negative | Neurological conditions (dementia;<br>other outcomes not analyzed by age<br>strata) |

| **Study**<br>**Country**<br>**Data source** | **Index time**<br>**for SARS-**<br>**CoV-2**<br>**infections** | **Study design** | **Mean/ median**<br>**follow-up, range**<br>**If not reported,**<br>**maximum**<br>**range of FU?** | **Care**<br>**type/setting**<br>**for SARS-**<br>**CoV-2**<br>**infected**<br>**cases** | **Age range**<br>**(years)**<br>**% Female** | **% Hospitalized**<br>**% in ICU** | **Comparator timing**<br>**Test status of**<br>**comparator group** | **Outcomes** |
| --- | --- | --- | --- | --- | --- | --- | --- | --- |
| Wang 2022a [49]<br>United States<br>TriNetX COVID-19<br>Research Network | Feb 2020 to<br>May 2021 | Retrospective<br>cohort | NR<br>Maximum of 360<br>days | Mixed | 65 to 85+<br>57% | NR | Concurrent<br>No<br>confirmed/suspected<br>SARS-CoV-2<br>infection | Neurological conditions<br>(Alzheimer's disease) |
| Wang 2022b [50]<br>United States<br>TriNetX COVID-19<br>Research Network | Jan 2019 to<br>Mar 2022 | Retrospective<br>cohort | NR<br>Minimum of 30<br>days to maximum<br>of 12 mos | Mixed | 20 to 65+<br>54% | NR | Concurrent<br>No<br>confirmed/suspected<br>SARS-CoV-2<br>infection | Cardiovascular disease (acute<br>coronary disease, angina, atrial<br>fibrillation and flutter, bradycardia,<br>cardiac arrest, cardiogenic shock,<br>cardiomyopathy, heart failure,<br>ischemic cardiomyopathy,<br>myocardial infarction, tachycardia,<br>ventricular arrhythmias); Stroke |
| Westman 2022 [51]<br>Sweden<br>SmiNET, Swedish<br>National Patient<br>Register | Feb 2020 to<br>Dec 2021 | Prospective<br>cohort | NR<br>Maximum of 22<br>mos | Mixed | 21 to 100+<br>51% | NR | Historical<br>NA | Neurological conditions (epilepsy) |
| Xie 2022a [52]<br>United States<br>Department of<br>Veterans Health<br>Administration | Mar 2020 to<br>Sept 2021 | Prospective<br>cohort | Cov: Median<br>(IQR) – 352 (244 –<br>406) days<br>Con: Median<br>(IQR) – 352 (245 –<br>406) days | Mixed | 0 to 65+<br>11.5% | 8.3<br>2.2 | Concurrent<br>No<br>confirmed/suspected<br>SARS-CoV-2<br>infection | Diabetes |
| Xie 2022b [53]<br>United States<br>Department of<br>Veterans Health<br>Administration | Mar 2020 to<br>Jan 2021 | Prospective<br>cohort | Cov: Median<br>(IQR) – 347 (317 –<br>440) days<br>Con: Median<br>(IQR) – 348 (318 –<br>441) days | Mixed | 0 to 65+<br>10% | 10.9<br>3.5 | Concurrent<br>No<br>confirmed/suspected<br>SARS-CoV-2<br>infection | Cardiovascular disease (dysrhythmia,<br>ischemic heart disease); Stroke |

| Study<br>Country<br>Data source | Index time<br>for SARS-CoV-2<br>infections | Study design | Mean/ median<br>follow-up, range<br>If not reported,<br>maximum<br>range of FU? | Care<br>type/setting<br>for SARS-CoV-2<br>infected<br>cases | Age range<br>(years)<br>% Female | %<br>Hospitalize<br>d<br>% in ICU | Comparator timing<br>Test status of<br>comparator group | Outcomes |
| --- | --- | --- | --- | --- | --- | --- | --- | --- |
| Xu 2022 [54]<br>United States<br>Department of<br>Veterans Health<br>Administration | Mar 2020 to<br>Jan 2021 | Prospective<br>cohort | Cov: Median<br>(IQR) – 408 (378 –<br>500) days<br>Con: Median<br>(IQR) – 348 (318 –<br>441) days | Mixed | 0 to 65+<br>10% | 10.8<br>3.4 | Concurrent<br>No<br>confirmed/suspected<br>SARS-CoV-2<br>infection | Mental disorders (anxiety disorders,<br>major depressive disorders,<br>psychotic disorders,<br>stress/adjustment disorders);<br>Neurological conditions (Alzheimer's<br>disease, memory problems) |
| Zarifkar 2022 [55]<br>Denmark<br>Electronic health<br>records from Capital<br>Region and Region<br>Zealand | Feb 2020 to<br>Nov 2021 | Prospective<br>cohort | NR<br>Maximum of 12<br>mos | Mixed | 18 to 80+<br>Inpatients: 51%<br>female<br>Outpatients:40%<br>females | 18.5<br>NR | Concurrent<br>>90% had test<br>negative | Neurological conditions<br>(Alzheimer's disease, Guillain-Barre<br>syndrome, multiple sclerosis,<br>myasthenia gravis, Parkinson's<br>disease); Stroke |

**Abbreviations:** Con = control group; Cov = SARS-CoV-2 infected group; FU = follow-up; ICU = intensive care unit; IQR = interquartile range; mos = months; NA = not applicable; NR = not reported; SD = standard deviation

To be eligible, studies had to report on severity of SARS-CoV-2 infection (i.e., hospitalization status), adjust for possible confounding by at least sex and two or more comorbidities (i.e., by matching, propensity scores, or multivariable regression), and report outcomes by age to allow for allocation to the most appropriate age group for analysis and synthesis of findings by key life-course stages: 0-17y, 18-64 y, and ≥65y. Study outcomes reported using differing age groupings were analyzed within the most appropriate age group.

Where a study reported an age group that spanned two of our categories, we weighted the data based on the number of years contributed to the age category. For example, data reported for 60-69 year-olds contributed to both the 18-64 year-old and ≥65 year-old groups but was given half of the 60-69 year-old age group’s overall weight. We included studies comparing people with confirmed (e.g., by laboratory testing) or suspected (e.g., physician diagnosed, regardless of test status) SARS-CoV-2 infection to those without. To ensure we would have some relevant studies to include, we did not require control groups to test negative for SARS-CoV-2. There was also no requirement for control groups to be healthy individuals (i.e., they could include hospitalized patients or individuals with other respiratory infections such as influenza but without SARS-CoV-2), to control for possible confounding, such as due to hospitalization not specific to SARS-CoV-2 infection.

Primary outcomes of interest were incidence and exacerbations of chronic conditions after SARS-CoV-2 infection compared to controls. Conditions of interest fell into the following categories: cardiovascular diseases, neurological conditions, cancer, chronic kidney disease, diabetes (excluding gestational diabetes), musculoskeletal disorders (e.g. osteoarthritis, gout, etc.), respiratory diseases, mental disorders, and stroke. Individual conditions within each category were also evaluated. Because of the limited clinical and epidemiologic relevance, [16] we did not look at dementia/cognitive impairment outcomes in individuals <18 years. Outcomes could be ascertained at any time after the acute phase of infection (i.e., immediately after discharge in hospitalized patients and ≥4 weeks in outpatients) and no minimum follow-up time was required. We attempted to only include studies reporting on diagnoses of chronic conditions, defined as those that were at a minimum documented by a healthcare provider in medical records; however, there may not have been standard diagnostic testing performed in all cases. Variables of interest for subgroup analyses were time since infection, vaccination status, and different SARS-CoV-2 variants of concern.

### Search Strategy

An information specialist (MT) developed a search strategy combining concepts for SARS-CoV-2 infection, post-acute/follow-up, outcomes (e.g., incidence), and chronic conditions of interest using vocabulary and syntax specific to each database searched. The search strategy was peer-reviewed by a second research librarian using the PRESS 2015 checklist. [17] Searches were carried out on Oct 4, 2022 in Ovid MEDLINE® including Epub Ahead of Print, In-Process & Other Non-Indexed Citations, and EMBASE. Search results were limited to those on or after 1 Jan 2020, and filters were applied to remove case reports, commentaries, and conference abstracts. The full searches for MEDLINE and EMBASE are available in Appendix 2 in the Supplement. In addition to database searches, a review lead (LG or JP) screened the reference lists of included studies and pertinent systematic reviews identified during screening for potentially relevant studies. Screening of reference lists and systematic reviews was completed on November 7, 2022.

### Study Selection

Search results were uploaded to an EndNote library (v. 20.3, Clarivate Analytics, Philadelphia, PA) and deduplicated before screening. Unique records were then uploaded to DistillerSR (Evidence Partners, Ottawa, Canada) and screened in a two-stage process, first by title and abstract (screening) and then by full-text (selection). Using standardized forms, all reviewers involved in screening and selection (LG, JP, SS) piloted the screening form with a random sample of 200 records and piloted the selection form with 16 full-text records from the database searches. Screening and selection proceeded once sufficient agreement between reviewers was reached.

During screening, DistillerSR’s machine learning feature (DistillerAI) was enabled. DistillerAI learns from human reviewers’ inclusion decisions to assign a likelihood score (0- 1, with values closer to 1 indicating higher likelihood of inclusion) for each unscreened record and prioritizes the most relevant records for screening by the human reviewers (i.e., the most relevant records are screened first). [18] Further, when threshold likelihood score for inclusion is applied, DistillerAI can act as a second reviewer with high specificity and sensitivity. [19] Thus, two reviewers independently screened the first 50% of titles and abstracts, after which DistillerAI acted as a second reviewer with likelihood threshold of 0.7. All remaining records with a DistillerAI-assigned likelihood >0.7 proceeded to selection and the rest were manually screened by one human reviewer for final exclusion. After screening, attempts were made to retrieve the full texts of all potentially relevant records. Two reviewers independently reviewed all retrieved full-texts and came to consensus on inclusion, with adjudication by a review lead or other reviewer (e.g., statistician) when necessary.

### Data extraction and management

We developed standardized data extraction forms in Microsoft Office Excel (v. 2019, Microsoft Corporation, Redmond, WA) which were independently piloted by all reviewers involved in extraction (LG, JP, SS). Thereafter, one reviewer extracted data from the included studies, and a second reviewer independently verified results data for accuracy and completeness. Disagreements were resolved by discussion. When relevant findings were reported in figures, data was extracted using Web Plot Digitizer (https://automeris.io/WebPlotDigitizer/). We only recorded zero events of a condition when it was explicitly reported.

We extracted the following information from each study: study characteristics (i.e., author, year, country, funding source, location of registration/protocol, design), population characteristics (i.e., inclusion and exclusion criteria, sample size, population demographics (age, sex, ethnicity, relevant comorbidities), SARS-CoV-2 infection confirmation method and timing), care setting during acute phase (outpatient, inpatient, mixed out- and inpatients), comparator(s), length of follow-up, analysis details (i.e., variables considered in analysis), outcome details (i.e., methods of ascertainment), and findings. For each condition category and/or individual condition of interest we extracted both relative (i.e., incidence rate ratios [IRRs] or hazard ratios [HRs]) and per-group incidence rates or cumulative incidence, when available. If an adjusted incidence rate or cumulative incidence was not reported (but participants were matched by at least sex and comorbidities), we extracted the crude number of events and estimated the cumulative incidence based on the denominator for each group. When results were reported for multiple time points, we took the longest follow-up. We extracted outcome data even when it was not able to be meta-analyzed, for example if only a p-value between groups was reported, to help interpret data and document possible reporting biases. Adjusted findings (i.e., from the most adjusted model) were prioritized in all cases.

We extracted any within-study analyses by time since infection, SARS-CoV-2 vaccination status, and different SARS-CoV-2 variants of concern and synthesized these narratively. Data extracted for this review are available on reasonable request from the authors.

### Risk of Bias Assessment

To assess risk of bias of included studies we used the JBI critical appraisal checklist for cohort studies. [20] After piloting, a review lead (LG) assessed the risk of bias for each study and brought any questions or concerns about included studies to the review team for discussion and consensus. We specifically considered in our assessment the validity of SARS-CoV-2 infection confirmation, with laboratory confirmed (using RT-PCR or antigen test) based on medical records being low risk and all others having some concerns. We also had concerns when a prospective study did not censor control participants who contracted COVID-19 during the follow-up period. We assigned an overall risk of bias rating (low, moderate or high) based on the number of questions answered “No” for each study (0 for low, 1 for moderate, ≥2 for high). Final assessments were incorporated into our certainty of evidence assessments guided by the Grading of Recommendations, Assessment, Development and Evaluation (GRADE) approach (see below). [21]

### Data Synthesis

We conducted random-effects meta-analysis using inverse variance weighting in Review Manager (RevMan; v5.4, The Cochrane Collaboration, 2020) to estimate a pooled hazard ratio when two or more studies reported on a condition category, or individual condition, by age category and COVID-19 care setting (inpatient vs. outpatient/mixed). Because our analysis was based on planned sub-groups, we did not investigate further into potential sources of heterogeneity. Forest plots were generated in RevMan to visually display results of the meta-analyses. Data not appropriate for meta-analysis were synthesized narratively. For all meta-analyses, a relative effect of 0.75-1.25 was considered little-to-no association; 0.51-0.74 and 1.26 to 1.99 small-to-moderate association (decrease or increase, respectively), and ≤0.50 or ≥2.00 large association. All studies with useable data were included in the meta-analyses for each condition category or individual condition they reported on. Since we identified no eligible studies with data on exacerbations of pre-existing conditions and new diagnosis of a condition can only occur once, we considered reported hazard ratios and incidence rate ratios to be interchangeable. When only cumulative incidence or crude events were reported, we estimated the incidence rates for each group by dividing the number of events by the average follow-up period (in years) multiplied by the number of participants.

We conducted separate analyses for each of the following categories of chronic conditions: cardiovascular disease, neurological conditions, chronic kidney disease, diabetes, musculoskeletal disorders (e.g. osteoarthritis, gout, etc.), respiratory diseases, mental disorders, and stroke. Although cancer was also among our chronic conditions of interest, we did not identify any eligible studies reporting on this outcome. The disease leads helped to ensure conditions reported by each included study were appropriately categorized. We also analyzed individual conditions within each condition category (e.g., dementia/mild cognitive disorder within the category of neurological conditions) when there was condition-specific data and a sample size of >2000 in the SARS-CoV-2 infection group.

For studies that reported data for multiple diagnoses falling within the label of an individual condition (e.g., tachycardia and ventricular arrhythmia, which would both contribute to the condition labelled “arrhythmias”), we calculated an estimated average for the condition weighted by the inverse of the variance to give more weight to results with more reliable estimates. This process was also used when a study reported multiple individual conditions within a condition category, if the study did not report a suitable composite outcome for the condition category.

For all categories and individual conditions with low, moderate or high certainty of some direction of effect (i.e., small-to-moderate or large increase/decrease), we estimated the excess incidence in the SARs-CoV-2 group per 1000 people over 6 months . We used a hierarchy to identify the most relevant data to use for the control (non-SARS-CoV-2) event rate. If at least one study reported a composite outcome (e.g., any cardiovascular event) within a condition category, we used the study’s reported incidence for that composite outcome. When a condition category had no directly reported composite incidence, we looked at the individual conditions in that condition category. Where we considered conditions within a condition category to be mutually exclusive (broadly speaking), we took the sum of their incidence in the control group as an estimate of the control event rate. Where conditions within a condition category were not mutually exclusive, we used the individual condition with the highest incidence as a conservative estimate. When multiple studies reported a control event rate for a condition category or individual condition, we took an average weighted by sample size. We converted all control event rates to a standard 6-month period, which was most representative of follow-up duration in the included studies. For example, a 1-year incidence rate was divided by 2 to estimate the incidence over 6 months. We estimated excess incidence by subtracting the control event rate from the product of the control event rate and the relative effect.

Other than age and COVID-19 care setting, we did not conduct any quantitative subgroup or sensitivity analyses. However, we planned to narratively summarize any time-varying effects and any within-study sub-group analyses for different SARS-CoV-2 variants of concern or by SARS-CoV-2 vaccination status.

### Certainty of Evidence

Two reviewers reached consensus through discussion on the certainty about conclusions in relation to our thresholds of effect of the relative effects for each outcome, guided by GRADE. [21, 22] We started the evidence at high certainty [23] and down rated to lower levels (i.e., moderate, low, and very low certainty) based on study quality in five domains (i.e., risk of bias, indirectness, inconsistency, imprecision, reporting biases). For each domain, we rated down by 0, 1, or 2 levels depending on the seriousness of the concerns, i.e., how much the domain appeared to impact the conclusions. We used thresholds as the targets of our certainty: a relative effect of 0.75 to 1.25 was considered little-to-no association, 0.51 to 0.74 and 1.26 to 1.99 was considered small-to-moderate, and ≤0.50 or ≥2.00 large. For example, we did not rate down for risk of bias when both high and low risk of bias studies had estimates surpassing the threshold for magnitude of association. Similarly, we did not rate down if some of our concerns in one domain likely stemmed from another domain, for example, we did not rate down for inconsistency if differences in estimates across studies were judged to be primarily related to risk of bias. If only one or two conditions contributed to a condition category estimate, we considered this an indirectness concern. To assess reporting biases, we compared outcomes specified in each report’s

Methods section (or protocol if available) with the outcomes reported in the Results section. Outcomes in the results section that were not specified in the methods or specified in the methods but not reported in the results were considered concerns. We rated down for inconsistency/lack of consistency when there was a single study in the analysis, when there was concerning variation not accounted for in other domains in study estimates (in relation to our thresholds), or when a single study contributed >80% weight to an estimate. Finally, we rated down once or twice for imprecision when one or both of the ends, respectively, of the confidence interval extended across an effect threshold (i.e., from effect into little-to-no difference or vice versa). When considering imprecision, we made conclusions about the results for which we had the highest certainty; for example when a point estimate surpassed our threshold for a large association but with imprecision, we instead made conclusions about a small-to-moderate association without imprecision concerns.

## Results

The flow of records through the selection process is depicted in Figure 1, and Appendix 3 in the Supplement lists relevant studies that did not meet key eligibility criteria, with reasons for exclusion. After screening 4,648 unique database records and 24 records identified from other sources, we included 25 studies from six countries: United States (15), Germany (4), United Kingdom (3), Denmark (1), Korea (1), and Sweden (1). The included studies are summarized in Table 1. Eight (32%) of the included studies confirmed that the control group was negative for SARS-CoV-2 using laboratory testing. Only 2 (8%) eligible studies reported on chronic conditions after hospitalization with SARS-CoV-2. We did not identify any eligible studies reporting on cancer, osteoarthritis, or gout after SARS-CoV-2 infection, nor did we identify any eligible studies reporting exacerbations of pre-existing chronic conditions.

**Figure 1.**
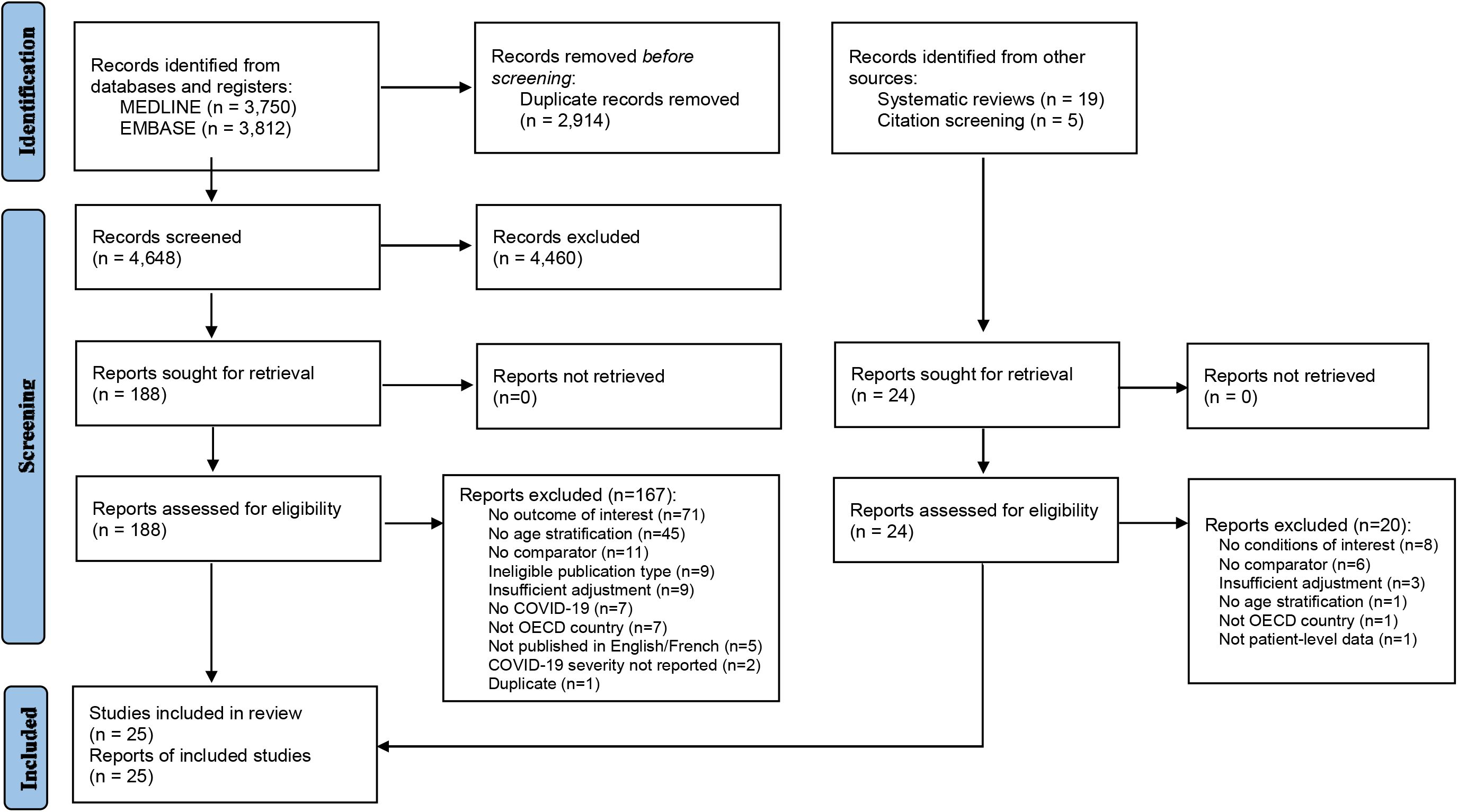
PRISMA flow diagram for a systematic review of the associations between SARS-CoV-2 infection and incidence of new chronic condition diagnoses. Template From: Page MJ, McKenzie JE, Bossuyt PM, Boutron I, Hoffmann TC, Mulrow CD, et al. The PRISMA 2020 statement: an updated guideline for reporting.

Risk of bias assessments are presented in Table 2. The majority of studies (18/25, 72%) were considered moderate risk of bias with only three having low risk. The most frequent concern for risk of bias was the potential for misclassification, largely due to differential exposure ascertainment methods between groups mostly from not confirming the absence of exposure with negative tests in the control group. Our assessment of potential reporting biases did not identify evidence of missing outcome data in any of the studies included in the meta-analysis.

**Table 2.** Risk of bias assessment according to JBI’s Cohort Studies tool

| Study | Q1 | Q2 | Q3 | Q4 | Q5 | Q6 | Q7 | Q8 | Q9 | Q10 | Q11 | Overall |
| --- | --- | --- | --- | --- | --- | --- | --- | --- | --- | --- | --- | --- |
| Abel 2021 | Y | N | U | Y | Y | Y | Y | U | U | U | Y | Moderate |
| Ayoubkhani 2021 | N | N | Y | Y | Y | Y | Y | Y | Y | NA | Y | High |
| Bohlken 2022 | Y | N | N | Y | Y | Y | Y | Y | Y | NA | Y | High |
| Chevinsky 2021 | U | U | Y | Y | Y | Y | Y | N | U | U | U | Moderate |
| Cohen 2022 | Y | N | Y | Y | Y | Y | Y | Y | Y | NA | Y | Moderate |
| Daugherty 2021 | Y | N | Y | Y | Y | Y | Y | Y | Y | Y | Y | Moderate |
| Donnachie 2022 | Y | Y | Y | Y | Y | Y | Y | Y | N | Y | N | High |
| Jacob 2022 | Y | N | Y | Y | U | Y | Y | U | U | Y | U | Moderate |
| Kompaniyets 2022 | Y | N | Y | Y | U | Y | Y | Y | U | Y | Y | Moderate |
| Park 2021 | Y | N | U | Y | Y | Y | Y | Y | Y | NA | Y | Moderate |
| Pietropaolo 2022 | Y | N | Y | Y | Y | Y | U | Y | U | U | Y | Moderate |
| Qureshi 2022 | Y | Y | Y | Y | Y | Y | Y | Y | U | U | U | Moderate |
| Rao 2022 | Y | Y | Y | Y | Y | Y | Y | Y | U | U | Y | Low |
| Rezel-Potts 2022 | Y | N | U | Y | Y | Y | Y | Y | Y | NA | Y | Moderate |
| Roessler 2022 | Y | N | U | Y | Y | Y | U | Y | U | Y | U | Moderate |
| Taquet 2022 | Y | N | Y | Y | Y | Y | Y | Y | Y | U | Y | Moderate |
| Taquet 2021 | Y | N | U | Y | Y | Y | Y | N | U | U | Y | High |
| Tartof 2022 | Y | Y | U | Y | Y | U | Y | Y | Y | NA | Y | Low |
| Wang 2022a | Y | N | Y | Y | Y | Y | Y | Y | U | U | Y | Moderate |
| Wang 2022b | Y | Y | Y | Y | Y | Y | Y | Y | Y | NA | Y | Low |
| Westman 2022 | Y | N | Y | Y | Y | Y | Y | U | Y | NA | Y | Moderate |
| Xie 2022a | Y | N | Y | Y | Y | Y | Y | U | Y | NA | Y | Moderate |
| Xie 2022b | Y | N | Y | Y | Y | Y | Y | U | U | U | Y | Moderate |
| Xu 2022 | Y | N | Y | Y | Y | Y | Y | U | U | U | Y | Moderate |
| Zarifkar 2022 | Y | Y | Y | Y | U | U | Y | Y | U | U | U | Moderate |

**Abbreviations:** N = no; NA = not applicable; U = unsure; Y = yes

Appendix 4 in the Supplement contains all forest plots. Table 3 presents the summary of findings, including the certainty of evidence for the relative effects and estimates of the excess cumulative incidence in 1000 people over 6 months (for outcomes with low, moderate or high certainty of a direction of effect). The GRADE domain(s) that led to rating down our certainty are documented in the table footnotes. One study included in our review reported on 31 conditions in 7 categories, but only provided non-stratified numeric results for adults (≥18y; ∼15% of sample was ≥65 y). This study reported results for children (<18 y) as a broad statement of no difference without effect estimates or variance, and thus was unable to be included in the meta-analysis. We do not report directions of effect for outcomes in which we had very low certainty. The condition categories and individual conditions for which we had moderate or high certainty were limited to the outpatient/mixed care group and are outlined below.

**Table 3.**
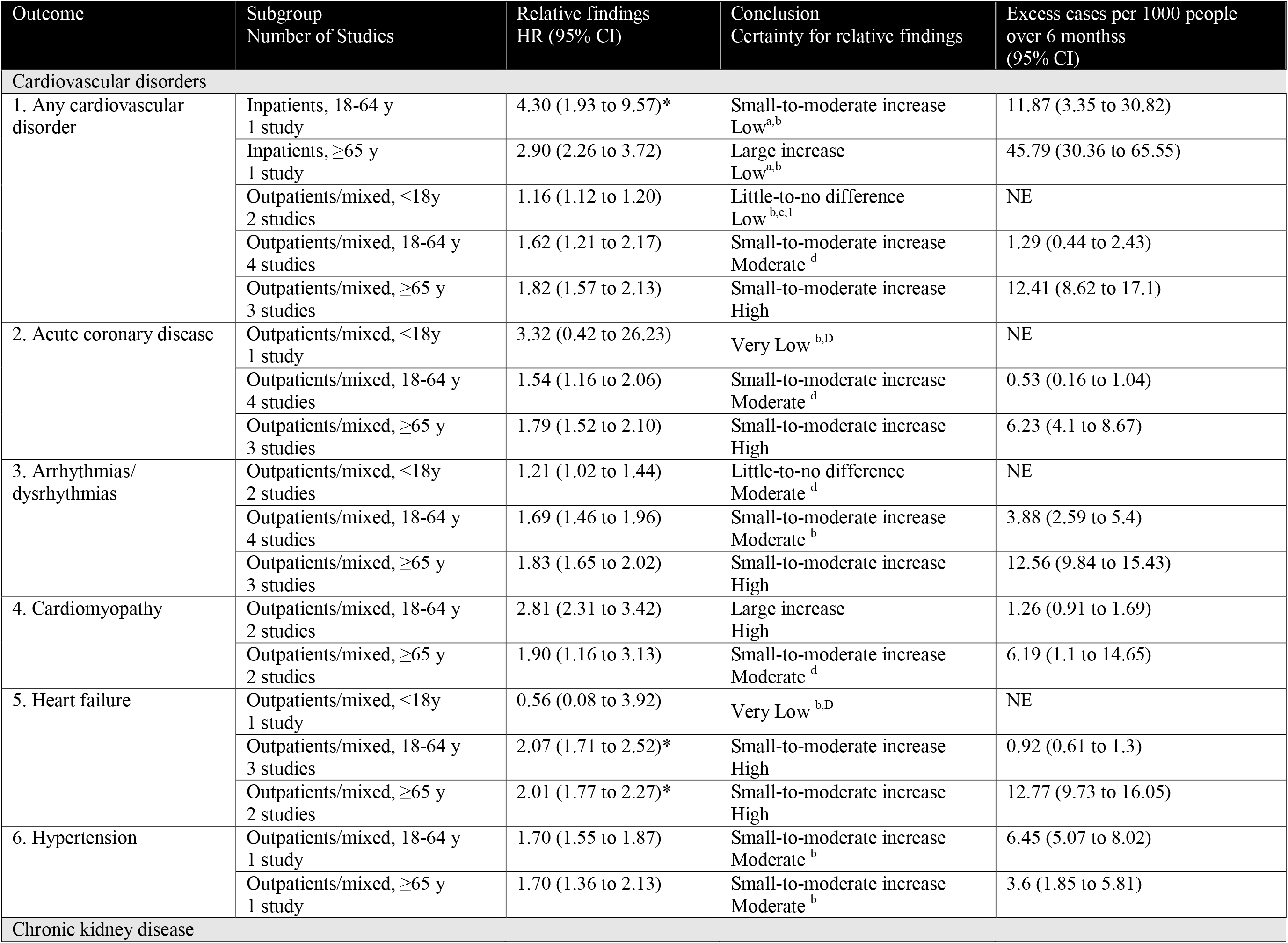
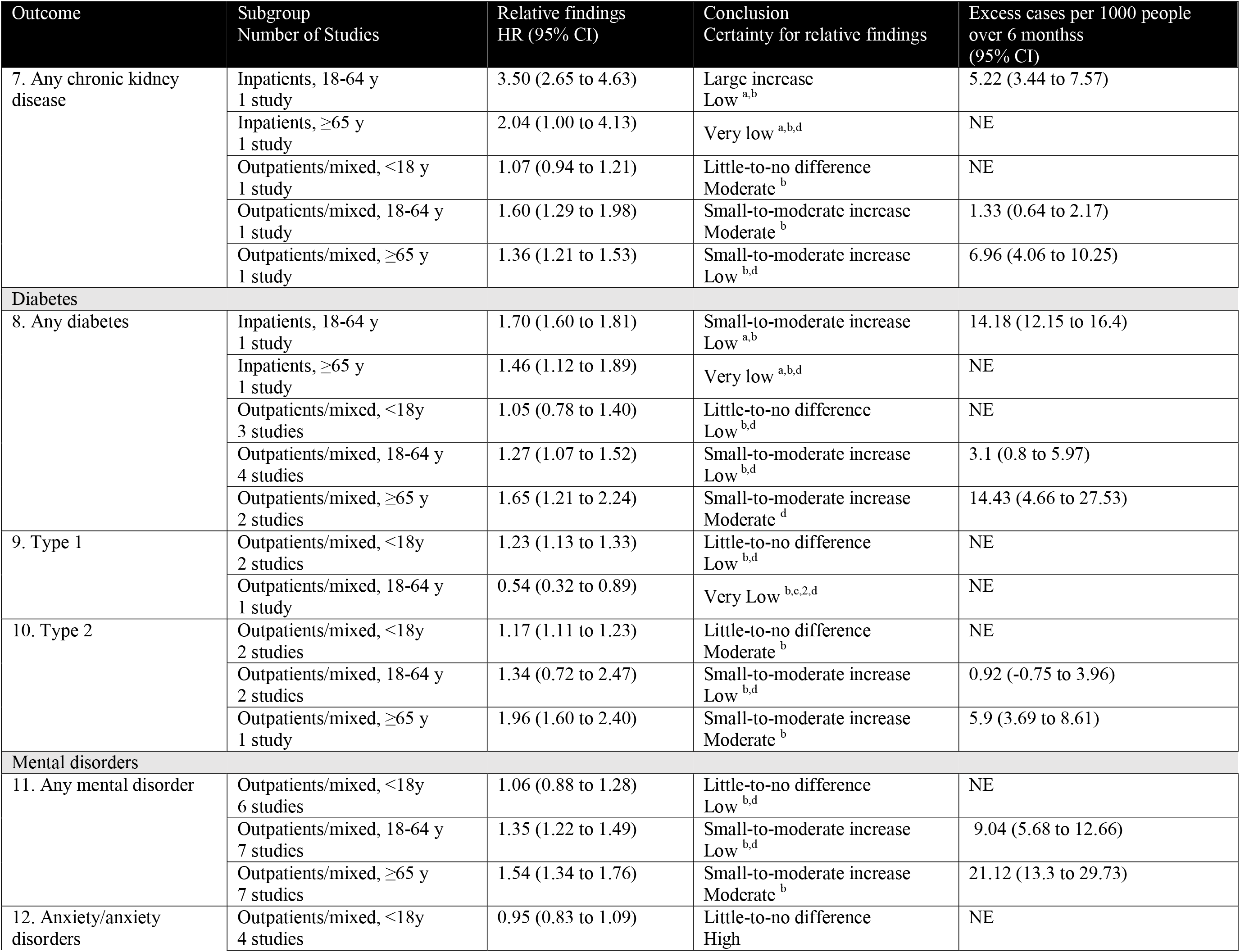
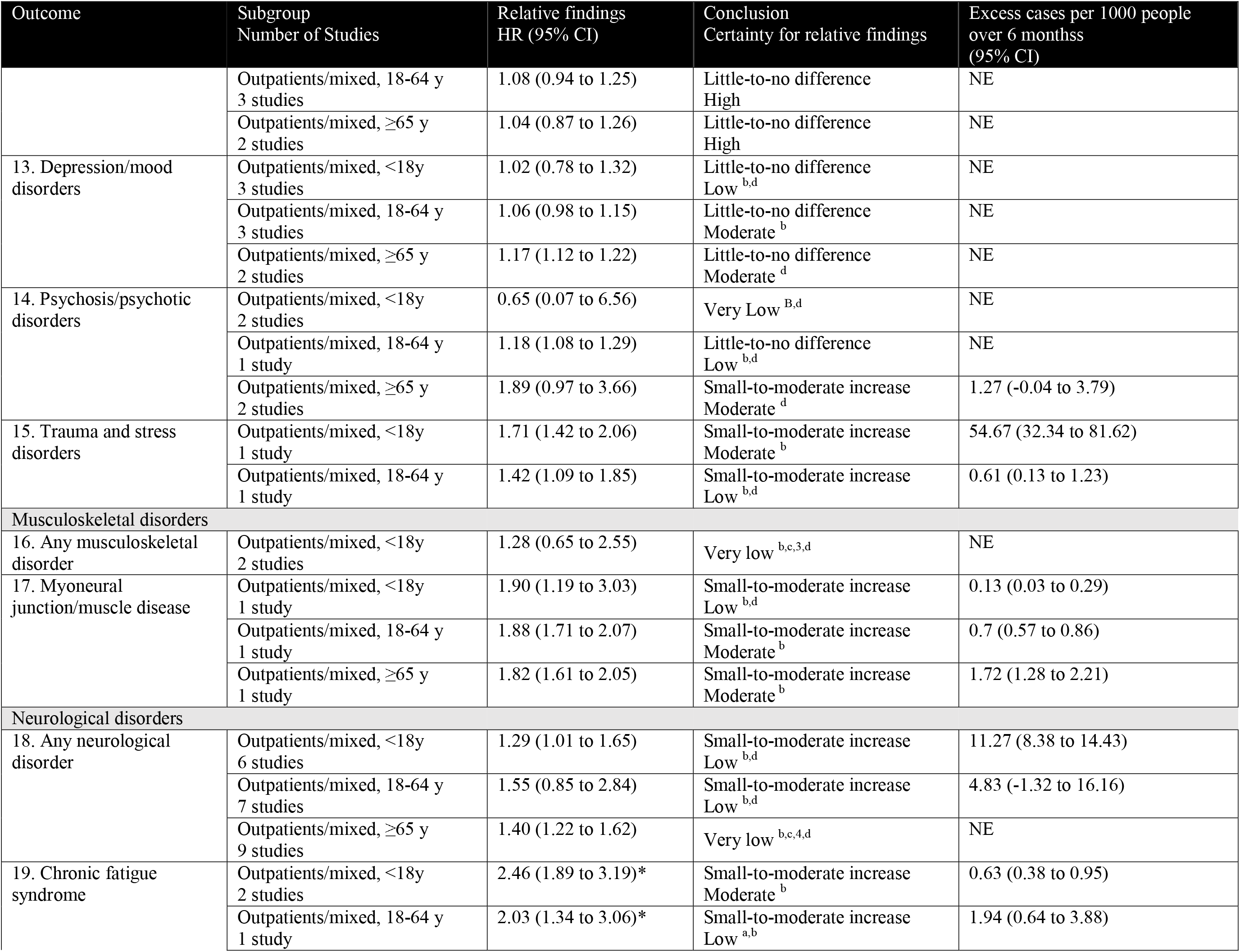
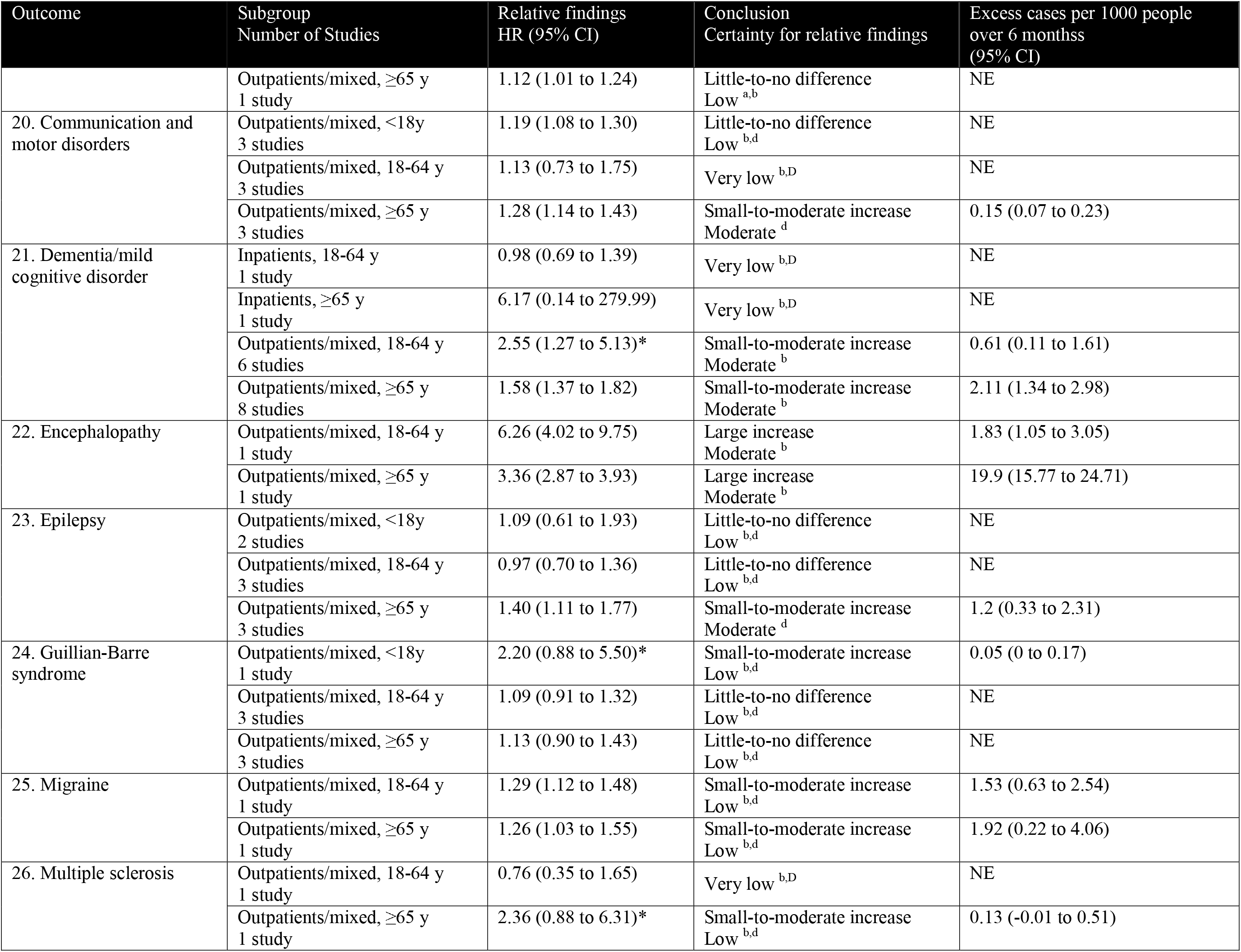
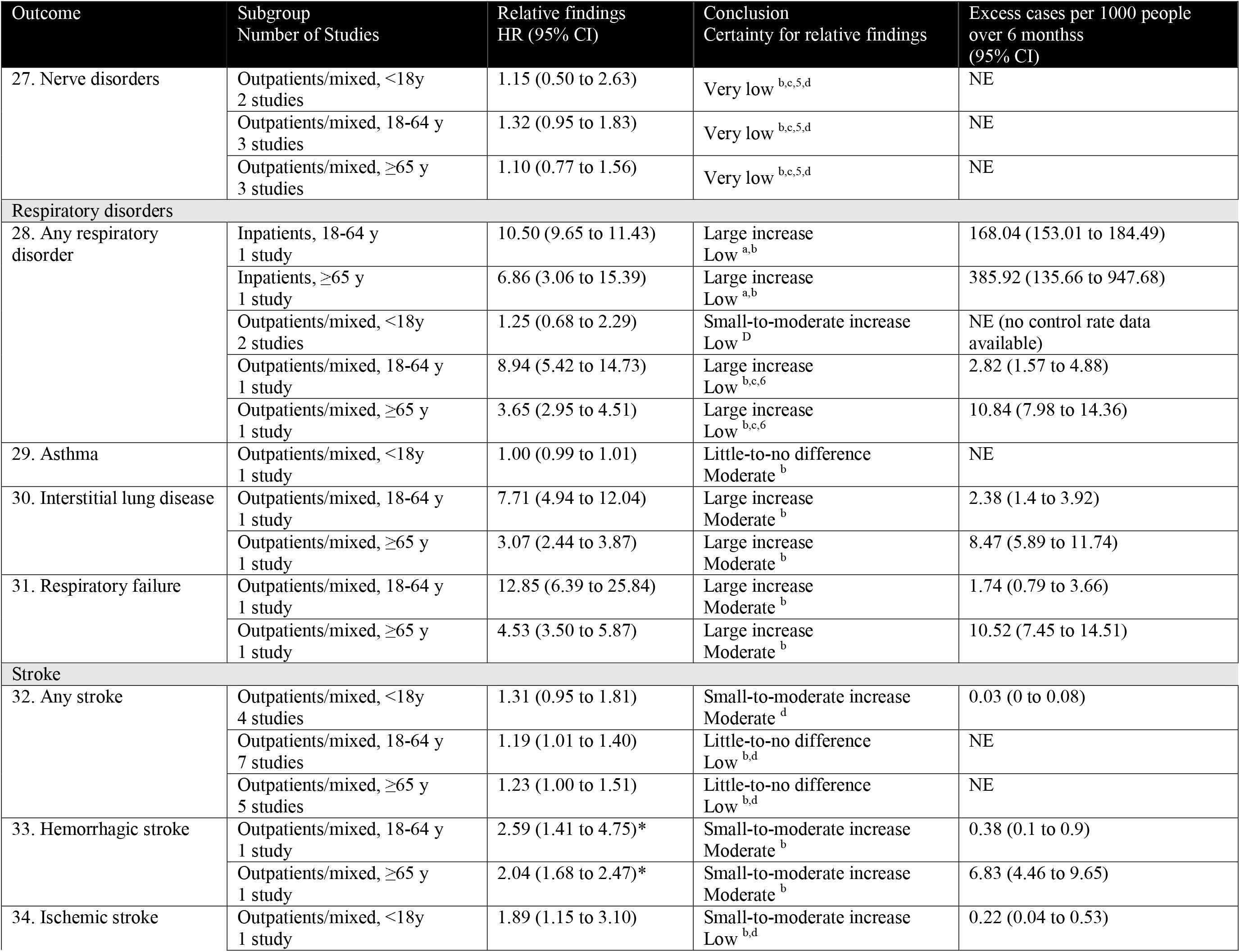
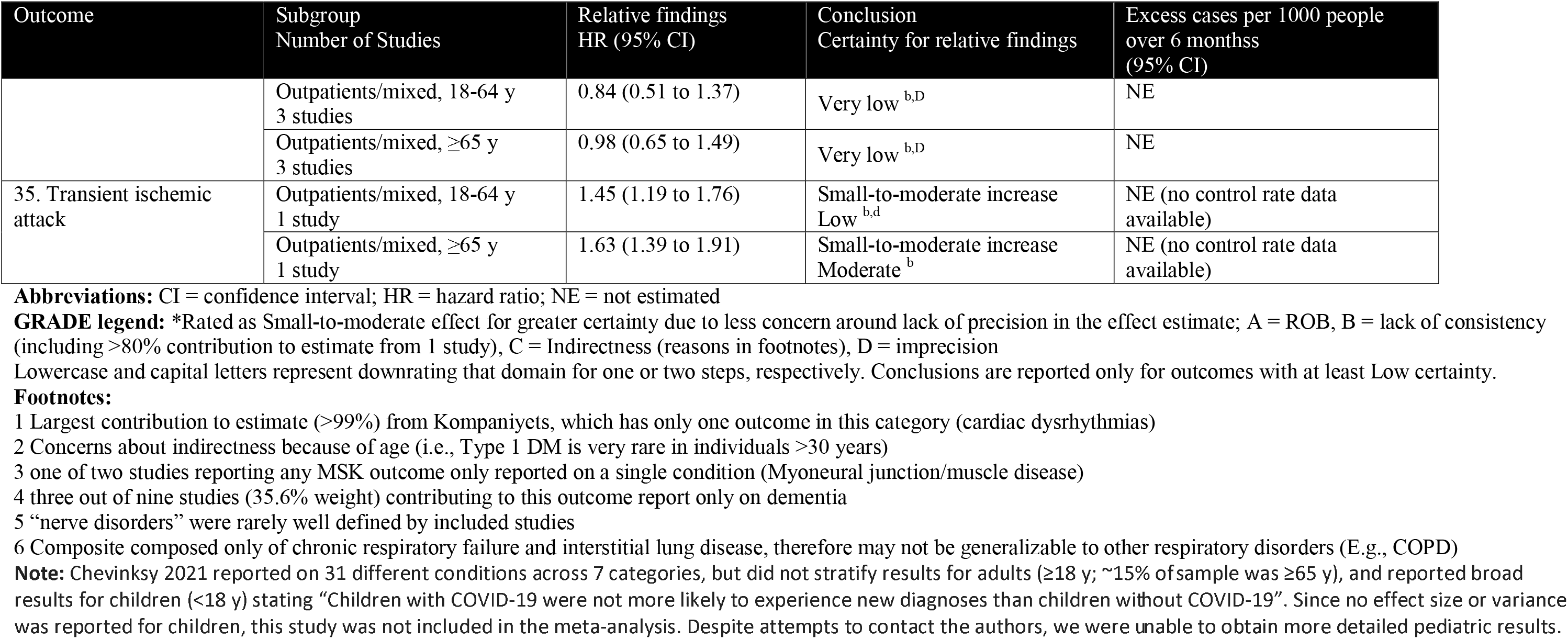
Summary of Findings for new diagnoses of chronic conditions after SARS-CoV-2 infection.

For ≥65 year-olds, we have high certainty that SARS-CoV-2 infection is associated with a small-to-moderate increase of any cardiovascular disorder, acute coronary disease, arrhythmias/dysrhythmias, and heart failure. For 18-64 year-olds, we have high certainty of a large increase in cardiomyopathy and of a small-to-moderate increase of heart failure. For all age groups (<18, 18-64, ≥65), we have high certainty of little-to-no difference in anxiety/anxiety disorders.

For individuals <18 years old, we have moderate certainty of a small-to-moderate increase in trauma and stress disorders, chronic fatigue syndrome, and stroke; and moderate certainty of little-to-no difference in arrhythmias/dysrhythmias, chronic kidney disease, type 2 diabetes, and asthma.

For 18-64 year-olds, we have moderate certainty of a large increase in encephalopathy, interstitial lung disease, and respiratory failure; moderate certainty of a small-to-moderate increase of any cardiovascular disorder, acute coronary disease, arrhythmias/dysrhythmias, hypertension, chronic kidney disease, myoneural junction/muscle disease, dementia/mild cognitive disorder, and hemorrhagic stroke; and moderate certainty of little-to-no difference for depression/mood disorders.

Among individuals ≥65 years old, we have moderate certainty of a large increase of encephalopathy, interstitial lung disease, and respiratory failure; moderate certainty of a small-to-moderate increase of cardiomyopathy, hypertension, any diabetes, type 2 diabetes, any mental disorder, psychosis/psychotic disorders, myoneural junction/muscle disease, communication and motor disorders, dementia/mild cognitive disorder, epilepsy, hemorrhagic stroke, and transient ischemic attack; and, moderate certainty of little-to-no difference for depression/mood disorders.

Two studies reported on how associations varied across time since infection or by variant of concern (Table 4). Change in risk over time since SARS-CoV-2 infection likely differs between conditions; however, there is not enough evidence to draw condition-specific conclusions at this time. One study reported on differing risks across variants of concern which suggested that risks may differ across variants, but these differences may also be confounded by average severity of acute disease and mortality of each variant. We did not identify any studies eligible for our review that looked at differences in risk between vaccinated vs. unvaccinated groups.

**Table 4.** Summary of time-varying effects and subgroup analyses by variant in a systematic review of new diagnoses of chronic conditions after SARS-CoV-2 infection.

| Study | Age group | Conditions reported | Time varying effects | Effects across Variants |
| --- | --- | --- | --- | --- |
| **Rezel-Potts 2022** [45] | 18-64 y | Cardiovascular disease, diabetes | For both cardiovascular disease and diabetes, IRR was highest at 4-7 weeks, decreasing over time to IRR ~1 by 24 weeks. | Not reported |
| **Taquet 2022** [47] | <18 y, 18-64 y, ≥65 y | Many | Outcomes fell into three categories: 1) within 2 years, HRs have returned to baseline (e.g., mood disorder, anxiety disorder, and ischaemic stroke) and cumulative incidence equalizes between cohorts;<br>2) HRs have returned to baseline within 2 years but equal cumulative incidence was not reached (i.e., myoneural junction or muscle disease);<br>3) HRs remained greater than 1 at the end of the follow-up period and new diagnoses are being made more frequently after COVID-19 diagnosis than after a diagnosis of another respiratory infection up to 2 years after the index event (e.g., dementia, psychotic disorders, epilepsy). These different risk trajectories were broadly similar in children, adults, and older adults. | Alpha vs. Delta vs. Omicron<br>Risk profiles differed across variants. Alpha: 6-month HRs did not notably change from before to after emergence of Alpha.<br>Delta: Increased 6-month HRs of anxiety disorders, insomnia, cognitive deficit, epilepsy or seizures, and ischaemic strokes, but a lower risk of dementia, were observed in those diagnosed after the emergence of the delta variant compared to those diagnosed before. These risks were compounded by an increased risk of death.<br>Omicron: After Omicron, patients were at an increased risk (over 140 days of follow-up) of dementia, mood disorders, and nerve, nerve root, and plexus disorders, and at a broadly similar risk of most other outcomes. All risks were largely offset by a reduced risk of death after the emergence of omicron.<br>The authors concluded: “The decreased composite risks of death and neurological or psychiatric sequelae are reassuring for patients. However, the ongoing risk of individual outcomes indicates that health services will likely continue to face a similar rate of these post-COVID-19 diagnoses even with SARS-CoV-2 variants that lead to otherwise less severe disease.” |

**Abbreviations:** HR = hazard ratio; IRR = incidence rate ratio

## Discussion

We conducted a systematic review to identify associations between SARS-CoV-2 infection and exacerbations of pre-existing or new diagnoses of chronic conditions. We stratified analyses by age category and severity of SARS-CoV-2 infection (using hospitalization during acute phase of infection as a proxy) to enable meaningful interpretation of the findings and because these are strong predictors of severity of outcomes both in the acute [24] and recovery stages of COVID-19. [25] After SARS-CoV-2 infection, there is probably an increased risk of new diagnoses for some, but not all chronic conditions. In general, we had the most certainty in associations between SARS-CoV-2 infection and new diagnoses of chronic conditions, especially cardiac conditions, in outpatient/mixed cares amples aged ≥65 years. People in this age category are already at increased risk of many chronic conditions and are more susceptible to poor outcomes after SARS-CoV-2 infection. [24, 26] We also had moderate to high certainty in associations between SARS-CoV-2 infection and at least a small increase of new diagnoses of several chronic conditions in individuals 18-64 years old and a few chronic conditions in individuals <18 years (i.e., trauma and stress disorders, chronic fatigue syndrome, and stroke). We identified only two eligible studies reporting on associations between hospitalization with SARS-CoV-2 infection and new diagnosis of chronic conditions. While it is widely recognized that severity of initial SARS-CoV-2 infection leads to poorer long-term outcomes, [25] we were not able to draw conclusions in any age group regarding an association between SARS-CoV-2 infection and subsequent new diagnoses of chronic conditions among individuals hospitalized during the acute infection phase. Finally, although we did not identify any eligible studies reporting on exacerbations of pre-existing chronic conditions after SARS-CoV-2 infection, this does not preclude the existence of this relationship for some conditions.

While previous systematic reviews have reported on the incidence of newly diagnosed chronic conditions after SARS-CoV-2 infection or reported on associations with specific conditions, [7–12] this is the first systematic review we are aware of that reports on associations between SARS-CoV-2 infection and new diagnoses of a wide range of chronic conditions specifically by age group. Our strict eligibility criteria, including the requirement to account for sex and relevant comorbidities, also likely reduced the number of eligible studies at high risk of bias. Overall, our findings suggest that there is probably an increased risk of diagnoses for some – but not all – chronic conditions after SARS-CoV-2 infection. In general, cardiovascular diseases and respiratory conditions showed the most consistent effects across adult age categories and disease severities. These associations have implications for decision makers in both policy and healthcare systems at a time when healthcare systems are already under considerable strain As the number of individuals infected by SARS-CoV-2 increases, so too will the number of new diagnoses for chronic conditions, leading to increased health care utilization in the form of specialty care, follow-up with primary care providers and increasing medication and treatment costs at either the patient or system level.

One notable gap highlighted by our review is the lack of evidence around cancer diagnoses after SARS-CoV-2 infection. This is not surprising, as there has likely been insufficient time since the start of the pandemic for disease processes and diagnosis, and longitudinal studies of this association have already been proposed. [27] However, such studies will have to be conducted with careful considerations of the impacts of the pandemic apart from SARS-CoV-2 infection. Public health restrictions during the early waves of the pandemic created access barriers to cancer screening and diagnosis, creating a potential backlog of missed screenings. [28] This may have resulted in delayed diagnoses and therefore will need to be controlled for in the study design of any longitudinal studies examining cancer incidence after SARS-CoV-2 infection.

We also found very few eligible studies examining how the potential risk of being diagnosed with a new chronic condition changes over time since SARS-CoV-2 infection, as well as across different variants of concern. Based on human tissue cultures and animal models, SARS-CoV-2 variants may preferentially infect or replicate in different organ systems or tissues, [29–31] and thus may result in a changing constellation of new chronic disease diagnoses after SARS-CoV-2 infection.

### Limitations

As with any systematic review, our synthesis comes with some limitations. First, while we made attempts to limit study eligibility to only those reporting on conditions documented or diagnosed by a medical provider, for some conditions it was not always possible to differentiate between chronic disorders versus persistent post-COVID symptoms. Second, while most studies included in this review used International Classification of Diseases (ICD)-10 codes (or similar administrative coding systems) to define outcomes of interest, there was substantial variation across studies in which codes were used to define each condition. This likely contributed to the substantial heterogeneity in estimates for some conditions. Third, some of the chronic conditions of interest are much simpler to diagnose than others. For example, diagnosis of type 2 diabetes relies on empirical signs and biological markers that can be objectively measured and diagnosed by primary care providers, whereas it may take a longer time after initially seeking care to be diagnosed with conditions such as chronic fatigue syndrome or mood disorders because they are typically diagnosed by specialists that patients may or may not have access to. Fourth, although some of the included studies attempted to control for differences in care-seeking behavior between control and SARS-CoV-2 infected groups (e.g., by matching on index date and only including control participants with at least one health care contact after their determined index visit), we did not evaluate this potential confounder as part of our synthesis. Although we would not expect the ability to obtain a diagnosis to differ between infected and non-infected people who have sought care, there are likely differences in the number of health care contacts between the groups. Thus, some of the associations identified in our review may be the result of surveillance biases. In other words, an undiagnosed chronic condition may have been present in some individuals prior to SARS-CoV-2 infection, but seeking care for the infection (and subsequent health care contacts for follow-up) resulted in the undiagnosed condition being diagnosed when it otherwise may not have been until further on in the disease progression.

Lastly, we used the event rates in non-SARS-CoV-2 infected control groups to estimate the excess incidence for conditions in which we had at least low certainty of some direction of effect, standardized to rates over six months; however, event rates were not always reported as 6-month rates. Our estimate of excess incidence assumes that the incidence of new diagnoses in the SARS-CoV-2 group is constant, e.g., that the rate is the same 2 months after infection as it is 6 months after infection, and there is no evidence that this assumption holds true.

## Conclusion

After SARS-CoV-2 infection, there is probably an increased risk of diagnoses for some, but not all, chronic conditions. However, the extent of increased risk that is directly caused by SARS-CoV-2 is uncertain due to other factors, such as increased health care contacts or monitoring in infected individuals, which are difficult to fully account for in observational study designs. Although the findings of this review likely apply well to the pandemic period, reflecting the pandemic’s current impact on healthcare availability and people infected by the virus, it is uncertain whether the impact will remain stable into future years. Finally, how this risk changes over time since infection or by variant of concern is uncertain.

## Data Availability

Data extracted for this review is available from the authors upon reasonable request.

## Acknowledgements

Thank you to Becky Skidmore for peer-reviewing the database search strategies. We also thank Jingxuan Zhang, Murray Weeks, and Cynthia Robitaille for their guidance and insightful comments on this manuscript.

## Funding Statement

The Public Health Agency of Canada funded this work under Contract no. 4500429095. The analyses, conclusions, opinions and statements expressed herein are solely those of the authors. No endorsement by the Public Health Agency of Canada is intended or inferred. Dr. Hartling is supported by a Canada Research Chair in Knowledge Synthesis and Translation, and is a Distinguished Researcher with the Stollery Science Lab supported by the Stollery Children’s Hospital Foundation.

## Declaration of Interests

The authors have no conflicts of interest to disclose.

## Notes

### Competing Interest Statement

The authors have declared no competing interest.

### Funding Statement

The Public Health Agency of Canada funded this work under Contract no. 4500429095. Dr. Hartling is supported by a Canada Research Chair in Knowledge Synthesis and Translation and is a Distinguished Researcher with the Stollery Science Lab supported by the Stollery Childrens Hospital Foundation.

