## Supplementary files for "Associations between SARS-CoV-2 infection and incidence of new chronic condition diagnoses: a systematic review"

### Supplement

#### Table S1. Eligibility criteria for a systematic review on the risk of new diagnoses of chronic conditions after SARS-CoV-2 infection

|  | Inclusion criteria | Exclusion criteria |
| --- | --- | --- |
| Population | People of any age and health status | None |
| Exposure | SARS-CoV-2 infection  Studies of people with confirmed (e.g., by laboratory testing) or suspected (e.g., physician diagnosed or self-reported but without a positive test) SARS-CoV-2 infection are eligible | Lack of confirmed or suspected SARS-CoV-2 infection |
| Comparator | People without confirmed or suspected SARS-CoV-2 infection. There is no requirement for people in the control group to have had a negative test. Controls do not need to be from a healthy population; e.g., hospitalized patients or individuals with respiratory infections other than SARS-CoV-2 infection may serve as controls. | No eligible control group |
| Outcomes | Any one of the four outcomes below, reported in age categories that allow for analysis in subgroups of <18 y, 18-65 y, and >65 y, and adjusted for sex and at least 2 (relevant) comorbidities:  1. Incidence rate ratio for new diagnosis of chronic condition of interest*  2. Excess incidence for new diagnosis of chronic condition  3. Relative risk of having an exacerbation/worsening of a previously diagnosed chronic condition  4. Excess rate of having an exacerbation/worsening of a previously diagnosed chronic condition  *Conditions of interest:   - cancer - cardiovascular disease (including myocardial infarction) - chronic kidney disease - diabetes - mental disorders - musculoskeletal disorders (e.g. osteoarthritis, gout, etc.) - neurological conditions - respiratory diseases - stroke | Outcomes not adjusted for sex and at least 2 comorbidities  Outcomes not reported by age categories allowing for sub-group analysis by <18 y, 18-65 y, or >65 y.  Excluded conditions:   - Gestational diabetes - Composite outcomes that include condition-specific or all-cause mortality - Anxiety or depression symptoms attributed to post-COVID condition |
| Timing | Outcomes ascertained at any time after the acute phase of infection (i.e., after discharge in hospitalized patients and ≥4 weeks in outpatients). | Outcomes ascertained only during acute phase of the infection |
| Study design | Prospective and retrospective observational studies using participant-level data (including control arms of RCTs, if otherwise eligible) | - Case series - Case reports - Systematic reviews - Editorials, commentaries, other opinion pieces |
| Language of full-text | English or French | Full-texts published in any language other than English or French |
| Dates of publication | 1 Jan 2020 or later | Dec 31 2019 or earlier |

#### Appendix 1. PRISMA 2020 Checklist

| **Section and Topic** | **Item #** | **Checklist item** | **Location where item is reported** |
| --- | --- | --- | --- |
| **TITLE** | | |  |
| Title | 1 | Identify the report as a systematic review. | Title page (Page 1) |
| **ABSTRACT** | | |  |
| Abstract | 2 | See the PRISMA 2020 for Abstracts checklist. | Page 2 |
| **INTRODUCTION** | | |  |
| Rationale | 3 | Describe the rationale for the review in the context of existing knowledge. | Page 3 |
| Objectives | 4 | Provide an explicit statement of the objective(s) or question(s) the review addresses. | Page 3 |
| **METHODS** | | |  |
| Eligibility criteria | 5 | Specify the inclusion and exclusion criteria for the review and how studies were grouped for the syntheses. | Page 4 and Table S1 |
| Information sources | 6 | Specify all databases, registers, websites, organisations, reference lists and other sources searched or consulted to identify studies. Specify the date when each source was last searched or consulted. | Page 6 |
| Search strategy | 7 | Present the full search strategies for all databases, registers and websites, including any filters and limits used. | Appendix 2 |
| Selection process | 8 | Specify the methods used to decide whether a study met the inclusion criteria of the review, including how many reviewers screened each record and each report retrieved, whether they worked independently, and if applicable, details of automation tools used in the process. | Page 6-7 |
| Data collection process | 9 | Specify the methods used to collect data from reports, including how many reviewers collected data from each report, whether they worked independently, any processes for obtaining or confirming data from study investigators, and if applicable, details of automation tools used in the process. | Page 7-8 |
| Data items | 10a | List and define all outcomes for which data were sought. Specify whether all results that were compatible with each outcome domain in each study were sought (e.g. for all measures, time points, analyses), and if not, the methods used to decide which results to collect. | Page 7-8 |
|  | 10b | List and define all other variables for which data were sought (e.g. participant and intervention characteristics, funding sources). Describe any assumptions made about any missing or unclear information. | Page 7-8 |
| Study risk of bias assessment | 11 | Specify the methods used to assess risk of bias in the included studies, including details of the tool(s) used, how many reviewers assessed each study and whether they worked independently, and if applicable, details of automation tools used in the process. | Page 8 |
| Effect measures | 12 | Specify for each outcome the effect measure(s) (e.g. risk ratio, mean difference) used in the synthesis or presentation of results. | Page 9 |
| Synthesis methods | 13a | Describe the processes used to decide which studies were eligible for each synthesis (e.g. tabulating the study intervention characteristics and comparing against the planned groups for each synthesis (item #5)). | Page 9 |
|  | 13b | Describe any methods required to prepare the data for presentation or synthesis, such as handling of missing summary statistics, or data conversions. | Page 9-10 |
|  | 13c | Describe any methods used to tabulate or visually display results of individual studies and syntheses. | Page 9 |
|  | 13d | Describe any methods used to synthesize results and provide a rationale for the choice(s). If meta-analysis was performed, describe the model(s), method(s) to identify the presence and extent of statistical heterogeneity, and software package(s) used. | Page 9-11 |
|  | 13e | Describe any methods used to explore possible causes of heterogeneity among study results (e.g. subgroup analysis, meta-regression). | Page 9 |
|  | 13f | Describe any sensitivity analyses conducted to assess robustness of the synthesized results. | Page 11 |
| Reporting bias assessment | 14 | Describe any methods used to assess risk of bias due to missing results in a synthesis (arising from reporting biases). | Page 11 |
| Certainty assessment | 15 | Describe any methods used to assess certainty (or confidence) in the body of evidence for an outcome. | Page 11 |
| **RESULTS** | | |  |
| Study selection | 16a | Describe the results of the search and selection process, from the number of records identified in the search to the number of studies included in the review, ideally using a flow diagram. | Figure 1 |
|  | 16b | Cite studies that might appear to meet the inclusion criteria, but which were excluded, and explain why they were excluded. | Appendix 3 |
| Study characteristics | 17 | Cite each included study and present its characteristics. | Table 1 |
| Risk of bias in studies | 18 | Present assessments of risk of bias for each included study. | Table 2 |
| Results of individual studies | 19 | For all outcomes, present, for each study: (a) summary statistics for each group (where appropriate) and (b) an effect estimate and its precision (e.g. confidence/credible interval), ideally using structured tables or plots. | Appendix 4 (forest plots) |
| Results of syntheses | 20a | For each synthesis, briefly summarise the characteristics and risk of bias among contributing studies. | Tables 1&2 |
|  | 20b | Present results of all statistical syntheses conducted. If meta-analysis was done, present for each the summary estimate and its precision (e.g. confidence/credible interval) and measures of statistical heterogeneity. If comparing groups, describe the direction of the effect. | Table 3, Appendix 4 (forest plots) |
|  | 20c | Present results of all investigations of possible causes of heterogeneity among study results. | Table 3, Appendix 4 (forest plots) |
|  | 20d | Present results of all sensitivity analyses conducted to assess the robustness of the synthesized results. | N/A |
| Reporting biases | 21 | Present assessments of risk of bias due to missing results (arising from reporting biases) for each synthesis assessed. | Page 13 |
| Certainty of evidence | 22 | Present assessments of certainty (or confidence) in the body of evidence for each outcome assessed. | Table 3 |
| **DISCUSSION** | | |  |
| Discussion | 23a | Provide a general interpretation of the results in the context of other evidence. | Page 16 |
|  | 23b | Discuss any limitations of the evidence included in the review. | Page 18-19 |
|  | 23c | Discuss any limitations of the review processes used. | Page 18-19 |
|  | 23d | Discuss implications of the results for practice, policy, and future research. | Page 16-17 |
| **OTHER INFORMATION** | | |  |
| Registration and protocol | 24a | Provide registration information for the review, including register name and registration number, or state that the review was not registered. | Page 4 |
|  | 24b | Indicate where the review protocol can be accessed, or state that a protocol was not prepared. | Page 4 |
|  | 24c | Describe and explain any amendments to information provided at registration or in the protocol. | N/A |
| Support | 25 | Describe sources of financial or non-financial support for the review, and the role of the funders or sponsors in the review. | Page 4 |
| Competing interests | 26 | Declare any competing interests of review authors. | Page 4 |
| Availability of data, code and other materials | 27 | Report which of the following are publicly available and where they can be found: template data collection forms; data extracted from included studies; data used for all analyses; analytic code; any other materials used in the review. | Page 4 |

#### Appendix 2. Search strategies

##### MEDLINE Search strategy – After Covid: Health conditions development/exacerbation

After Concept

1 - draft - Covid chronic - After concept_MEDLINE_EMBASE

Conditions Concept

1 - draft - Covid chronic - chronic disease_MeSH_MEDLINE

1 - draft - Covid chronic - chronic disease_PHAC KW3_MEDLINE

Incidence/Prev/Risk Concept

1 - draft - Covid chronic - chronic disease_IPRE3_ MEDLINE

Study Design Filter

1 - draft - Covid chronic - study design filter_MEDLINE

Fluff Filter

1 - draft - Covid chronic - Animal and fluff filter_MEDLINE

Database(s): Ovid MEDLINE(R) ALL 1946 to October 03, 2022
Search Strategy: After Covid_Health Conditions_MEDLINE_4Oct2022

Run date: 4Oct2022

| # | Searches | Results | Annotations |
| --- | --- | --- | --- |
| 1 | ("after covid" or post-covid or postcovid or post-coronavir* or post-coronavir*).ti,ab. | 8730 |  |
| 2 | ((post-acute or postacute or "after acute" or "after discharge" or "after hospital discharge" or sequela* or post-infect* or post-viral or postviral or post-discharg* or postdischarge or survivor$1) adj5 (COVID or COVID-19 or COVID19 or coronavirus* or corona virus* or 2019-nCoV or 19nCoV or 2019nCoV or nCoV or n-CoV or SARS-CoV-2 or SARS-CoV2 or SARSCoV-2 or SARSCoV2 or 2019-novel CoV or Sars-coronavirus2 or novel CoV)).ti,ab. | 3650 |  |
| 3 | (("more than" or longer or "at least" or "after" or follow* or upward* or later* or prospectiv* or retrospectiv* or review* or longitud* or post or beyond) adj5 (day? or week? or wk or wks or month? or mo or mos or year? or yr or yrs)).ti,ab. | 2697628 |  |
| 4 | (COVID or COVID-19 or COVID19 or coronavirus* or corona virus* or 2019-nCoV or 19nCoV or 2019nCoV or nCoV or n-CoV or SARS-CoV-2 or SARS-CoV2 or SARSCoV-2 or SARSCoV2 or 2019-novel CoV or Sars-coronavirus2 or novel CoV).ti. | 262488 |  |
| 5 | 3 and 4 | 24827 |  |
| 6 | 1 or 2 or 5 | 32854 | After concept |
| 7 | Chronic Disease/ | 277651 |  |
| 8 | (Chronic adj3 (condition$1 or diagnos$2 or disease$1 or disorder* or failure or illness*)).ti,ab,kf. | 474693 |  |
| 9 | ((longterm or long-term) adj3 (consequence* or damage or effect$1 or impact* or outcome* or result* or sequela*)).ti,ab,kf. | 249921 |  |
| 10 | exp Cardiovascular Disease/ | 2652994 |  |
| 11 | exp myocardial revascularization/ | 96439 |  |
| 12 | exp Dementia/ | 195989 |  |
| 13 | diabetes mellitus, type 1/ or diabetes mellitus, type 2/ or diabetic ketoacidosis/ or prediabetic state/ | 239355 |  |
| 14 | exp Gout/ | 13568 |  |
| 15 | joint diseases/ or rheumatic diseases/ | 49241 |  |
| 16 | exp Osteoarthritis/ | 74173 |  |
| 17 | exp Mental Disorders/ | 1393816 |  |
| 18 | exp Neoplasms/ | 3744123 |  |
| 19 | exp Nervous System Diseases/ | 2788526 |  |
| 20 | exp Pulmonary Disease, Chronic Obstructive/ | 64629 |  |
| 21 | exp Renal Insufficiency, Chronic/ | 130404 |  |
| 22 | ("cardiovascular stroke*" or "heart attack*" or "heart infarct*" or "myocardial infarct*").ti,ab. | 220528 |  |
| 23 | "myocardial revasculari?ation".ti,ab. | 5014 |  |
| 24 | (((cardio* or cardia* or heart? or myocardi* or vascula* or atria* or ventric* or coronary) and (disease* or disorder* or condition* or syndrome* or illness* or abnormal* or dysfunction* or disturb* or event* or adverse* or inflam*)) or nstemi or stemi or cvd or fibrillation or paroxysmal dyspnea or arrhythmia* or dysrhythmia* or bradycard* or brugada syndrome* or commotio cordis or long qt syndrome or parasystole or pre-excitation syndrome* or tachycardia* or conduction disturbance*).ti,ab. | 1627525 |  |
| 25 | ("cerebrovascular accident*" or stroke or strokes).ti,ab. | 299814 |  |
| 26 | (diabet* or "t1dm" or "t1d" or "t2dm" or "t2d" or "niddm" or insulin resistan* or prediabet* or dysglyc?emi* or hyperglyc?emi* or hypoglyc?emi* or hyper glyc?emi* or hypo glyc?emi* or "hyperosmolar state*" or ketoacidosis or (impaired fasting adj2 (glucose or sugar)) or ((glucose or carbohydrate*) adj3 (toler* or intoler*))).ti,ab. | 846121 |  |
| 27 | ((chronic obstructive adj4 (pulmonary or airway? or lung?)) or chronic airflow obstruction* or copd or coad or aecopd or obstructive pulmonary or asthma* or emphysema* or (chronic adj3 (bronchitis or bronchus infection* or bronchi infection* or bronchus inflammation or bronchi inflammation))).ti,ab. | 269568 |  |
| 28 | (osteopor* or arthrit* or antisynthetase syndrome or behcet* disease or blau syndrom* or monarthrit* or monoarthrit* or coxitis* or sacroiliitis or osteoarthrit* or arthrosis* or spondylosis* or barre criecou or barre lieou or brachialgia paraesthetica nocturna or brachialgia paresthetica nocturna or neri barre or osteophytosis or polyspondylitis or spondylarthrosis or spondyloarthrosis or papa syndrome or papga syndrome or pigmented villonodular synovitis or "pvns" or reiter syndrome* or arthragra* or gout* or cheiragra* or chiragra* or urate inflammation* or pseudogout* or (crystal* adj3 arthropath*)).ti,ab. | 379982 |  |
| 29 | (epilep* or seizure* or panayiotopoulos syndrome or rasmussen syndrome or alpers disease or hypsarrhythmia or infantie spasm* or lennox gastaut syndrome* or merrf syndrome* or myoclonus* or nodding syndrome* or lipofuscinosis or (sclerosis adj3 (multiple* or disseminate* or insular*)) or chariot disease* or parkinsonism* or hemiparkinsonism* or (parkinson* adj3 (syndrom* or disorder* or disease*)) or paralysis agitans or amnesia* or delirium or alzheimer* or aphasi* or diffuse neurofibrillary tangle* or lobar degeneration* or acquired dyslexia or alexia or cadasil or cada sil or cerebral autosomal dominant arteriopathy or lewy body disease* or (pick* adj1 (complex* or disease* or disorder* or syndrome*)) or huntington* or juvenile chorea or chorea major or kluver bucy or (mental* adj3 deteriorat*) or pseudodementia* or rett syndrome* or senile* or tauopath* or ((neuro* or neural* or cognitive* or cognition* or consciousness or sleep* or wake*) adj3 (impair* or side effect* or function* or disfunction* or dysfunction* or disorder* or degenerat* or disease*))).ti,ab. | 1114345 |  |
| 30 | (((chronic kidney or chronic renal) adj2 (disease? or insufficienc* or failur?)) or CKD or chronic nephropath*).ti,ab. | 100656 |  |
| 31 | (((cancer* or tumor? or tumour?) adj3 (induct* or form* or genesis or caus*)) or cancerogen* or neoplasmogenesis or oncogenesis or oncogenicit* or tumorigenicit* or tumorigenesis or tumourgenicit* or tumourigenesis or carcinogeni* or (cell adj3 proliferat*) or clonal expan* or (foci adj3 alter*) or (reactivat* adj4 dorman*) or adamantinoma* or adenoacanthoma* or adenocanthoma* or adenocarcinoma* or adenomatos* or adenomyoepithelioma* or adenomyoma* or adenosarcoma* or aesthesioneuroblastoma* or ameloblastoma* or androblastoma* or angioblastoma* or angioendothelioma* or angiofibrosarcoma* or angiosarcoma* or argentaffinoma* or arrhenoblastoma* or astroblastoma* or astrocytoma* or astroglioma* or baltoma or basiloma* or basalioma* or blastoma? or buschke-lowenstein or cancer* or carcinogen* or carcinoid? or carcinoma* or carcinosarcoma* or chloroma* or cholangiocarcinoma* or cholangiohepatoma* or cholangiosarcoma* or chondroblastoma* or chondrosarcoma* or chorioadenoma* or chorioangioma or choriocarcinoma* or chorioepithelioma* or chorionepithelioma* or comedocarcinoma* or cystadenocarcinoma* or cystosarcoma* or dermatofibrosarcoma* or dictyoma* or dysgerminoma* or ectomesenchymoma* or ependymoblastoma* or erythroleukaemia* or erythroleukemia* or erythroplakia* or esthesioneuroblastoma* or esthesioneuroepithelioma* or fibroadenosarcoma* or fibrochondrosarcoma* or fibroepithelioma* or fibroliposarcoma* or fibrosarcoma* or fibroxanthoma* or fibroxanthosarcoma* or ganglioblastoma* or ganglioneuroblastoma* or glioblastoma* or gliomatos* or gliosarcoma* or haemangioblastoma* or haemangiopericytoma* or haemangiosarcoma* or hemangioblastoma* or hemangioendothelioma* or hemangioendotheliosarcoma* or hemangiopericytoma* or hemangiosarcoma* or hepatoblastoma* or hepatocarcinoma* or hepatocholangiocarcinoma* or hepatoma? or hodgkin* or hypernephroma* or immunocytoma* or keratoacanthoma* or leiomyosarcoma* or leukaemia* or leukemia* or liposarcoma* or lymphoma? or lymphangiosarcoma* or lymphoepithelioma* or macroglobulinemia* or malignan* or maltoma* or medulloblastoma* or medullomyoblastoma* or melanoameloblastoma* or melanoma* or meningioblastoma* or mesenchymoma* or mesonephroma* or mesothelioma* or metasta* or microglioma* or myelodysplas* or myeloma* or myelomatos* or myeloproliferat* or myosarcoma* or myxoliposarcoma* or nephroblastoma* or neuroblastoma* or neurofibrosarcoma* or nonhodgkin* or nonseminoma* or oligodendroglioma* or oncocytoma* or oncogen* or oncolog* or orchioblastoma* or osteoblastoma* or osteochondrosarcoma* or osteofibrosarcoma* or osteosarcoma* or pancreatoblastoma* or paraganglioma* or paraneoplastic or pinealblastoma* or pinealoblastoma* or pineoblastoma* or pneumoblastoma* or polyembryoma* or polyhistioma* or porocarcinoma* or precancer* or preleukaemia* or preleukemia* or premalignant* or preneoplastic* or reticuloendothelioma* or retinoblastoma* or rhabdomyosarcomas* or rhabdosarcoma* or sarcoma* or seminoma* or somatostatinoma* or teratocarcinoma* or waldenstrom* or xanthosarcoma*).ti,ab. | 3960430 |  |
| 32 | (anxiety* or depression* or schizo* or psychotic* or psychosis or psychoses or amnesia* or delirium* or gender dysphoria* or paraphilia* or tourette* or hallucination* or paranoid or paranoia or paraphrenia or sexual dysfunction* or hysteria or phobia? or agoraphobia or neurasthenia or hypochondriasis or hypochondria or hypochondriac or adjustment reaction*).ti,ab. | 757985 |  |
| 33 | ((manic or mania or depress*) adj3 episode*).ti,ab. | 14906 |  |
| 34 | ((bipolar* or personality or mood* or conduct or impulse or dissociative or eating or neurotic or obsessive or compulsive or panic or phobic or neurocognitiv* or trauma* or posttrauma* or stress* or neurocognitive* or amnestic* or behaviour* or behavior* or neurobehav* or delusion* or manic or mania or depress* or conversion or somatoform or gender identity or attention-deficit or hyperactivity or conduct or emotion* or tic or tics or sleep or sexual identity or dysthymic or alcohol-induced or drug-induced or social functioning) adj3 disorder*).ti,ab. | 292177 |  |
| 35 | ((mental* or psychiatric* or psychological* or neuropsych*) adj3 (disorder* or ill or illness* or diagnosis or diagnoses)).ti,ab. | 181447 |  |
| 36 | ((intellectual* or developmental* or neurodevelopmental or learning) adj3 (disabilit* or disorder* or syndrome*)).ti,ab. | 67238 |  |
| 37 | (alcoholism* or ((glue or gas) adj3 (sniffing or huffing)) or ((nicotine or tobacco) adj3 (dependen* or abus* or misus* or addict* or overdos* or over dos* or "use disorder*"))).ti,ab. | 43060 |  |
| 38 | ((drug? or substance? or multidrug? or polydrug? or polysubstance? or stimulant? or alcohol or sedative* or hypnotic* or anxiolytic* or psychoactive*) adj4 (abus* or misus* or addict* or "use disorder*")).ti,ab. | 125553 |  |
| 39 | ((adderall or angel dust or amphetamine? or ayahuasca or barbiturate* or benzodiazepine* or buprenorphine or crack or cocaine or codeine or demerol or dextroamphetamine or dextromethamphetamine or diamorphine or dihydromorphine or dilaudid or ecstasy or ethylmorphine or fentanyl or hallucinogen* or heroin or hydrocodeine or hydromorphone or hyrdocodone or ketamine or kratom or lsd or lysergic acid diethylamide or magic mint or mdma or meperidine or mescaline or methadone or meth or methamphetamine or methylenedioxyamphetamine or methylenedioxymethamphetamine or methylphenidate or methylphenidate or midomafetamine or molly or naltrexone or pcp or pethidine or peyote or phencyclidine or psilocybin or ritalin or salvia or suboxone or tramadol or ghb or gamma?hydroxybutyrate or inhalent? or inhalant? or narcotic? or opioid* or opiate* or opium or opon or dextropropoxyphene or anileridine or alphaprodine or diamorphine or levorphanol or sufentanil or bromadol or bdpc or etorphine or dihydroetorphine or carfentanil or heroin or alfentanil or remifentanil or etorphine or buprenorphine or anorfin or buprenex or buprex or buprine or butrans or norphin or pentorel or prefin or probuphine or subutex or temgesic or transtec or bupe or bute or suboxone or zubsolv or subby or sobos or codein? or codipertussin or codyl or methylmorfine or methylmorphine or pentuss or "tylenol 2" or "tylenol 3" or "tylenol 4" or cody or fentanyl or abstral or duragesic or durogesic or fentamyl or fentanil or fetnanyl or instanyl or ionsys or lazanda or leptanal or onsolis or pecfent or phentanyl or rapinyl or recuvyra or subsys or tanyl or transfenta or onsolis or bekadid or dico or dicodid or dihydrocodeinone or dihydrocodone or hydrocodone* or hydrocodonum or tussionex or vicoprofen or vike or hydromorphone or dilaudid or dillies or meperidine or pethidine or algil or alodan or centralgin or centralgine or cluyer or demero or demerol or dispadol or dolanquifa or dolantal or dolantin or dolantina or dolantine or dolargan or dolcontral or dolenal or dolestin or dolestine or dolin or dolocontral or doloneurin or doloneurotrat or dolor or dolosa or dolosal or dolosan or dolsin or dolvanol or endolate or isonipecaine or lidol or lydol or mefedina or mepadin or meperdol or meperiden or meperidin or meperidol or mephedine or mepiridine or mialgin or neomochin or opistan or pantalgin or petadin or petantin or petantina or pethanol or pethedine or pethidin or petidin or phetidine or piridosal or sauteralgyl or simesalgina or supplosal or synlaudine or demmies or methadone or adanon or algidon or algolysin or algoxale or althose or amidon or amidona or amidone or amidosan or anadon or biodone or butalgin or deamin or depridol or diaminon or dianone or dolafin or dolamid or dolesone or dolmed or dolophine or dorex or dorexol or eptadone or fenadon or gobbidona or heptadon or heptanon or ketalgin or mecodin or mepecton or mephenon or metadol or metadon or metasedin or methaddict or methadon or methadose or "methaforte mix" or miadone or moheptan or pallidone or phenadon or physepton or physeptone or polamidon or polamivet or polamivit or sinalgin or symoron or westadone or meth or morphine or anpec or duromorph or epimorph or miro or morfin or morfine or morphia or morphin or morphinium or morphium or opso or skenan or doloral or statex or "m.o.s." or morph or "red rockets" or oxycodone or bionine or bionone or bolodorm or broncodal or bucodal or cafacodal or cardanon or codenon or dihydrohydroxycodeinone or dihydrohydroxydodeinone or dihydrone or dinarkon or endone or eubine or eucodal or eucodale or eucodalum or eudin or eukdin or eukodal or eumorphal or eurodamine or eutagen or hydrocodal or hydroxycodeinoma or ludonal or "m-oxy" or medicodal or narcobasina or narcobasine or narcosin or nargenol or narodal or nucodan or opton or ossicodone or oxanest or oxecta or oxicone or oxicontin or oxiconum or oxikon or "oxy ir" or oxycod or oxycodeinonhydrochloride or oxycodyl or oxycone or oxycontin or oxydose or oxyfast or oxygesic or oxyir or oxykon or oxynorm or pancodine or pavinal or percolone or pronarcin or remoxy or roxicodone or roxycodone or sinthiodal or stupenal or supeudol or tebodal or tekodin or thecodin or oxyneo or percocet or oxycocet or percodan or oxy or percs or pentazocine or dolapent or fortal or fortalgesic or fortral or fortraline or fortwin or liticon or peltazon or pentacozine or pentafen or pentagin or pentalgina or pentazocin or pentozocine or perutagin or sosegon or sosigon or talioin or talwin or tapentadol or nucynta or palexia or yantil or nucynta or tramadol or adamon or amanda or analab or analdol or andalpha or bellatram or biodalgic or calmador or calmol or contramal or dolana or dolika or dolmal or dolotral or dolzam or dromadol or eufindol or exopen or katrasic or mabron or melanate or mosepan or newdorphin or nobligan or nonalges or omnidol or pengesic or penimadol or prontofort or radol or rofy or ryzolt or sefmal or sensitram or takadol or tamolan or tandol or tarol or topalgic or trabar or trabilan or trabilin or tradol or tradolan or tradonal or tralic or tramada or tramadex or tramagetic or tramagit or tramahexal or tramake or tramal or tramazac or tramed or tramol or tramundin or trasedal or trasik or "trd-contin" or trexol or tridol or trodon or trondon or ultram or unitral or urgendol or zamadol or zamudol or zodol or zumatran or zydol or tramacet or tridural or durela or "chill pill?" or oxymorphone or opana or algopan or quarelin or actiq or fentora or biopan or cofapon or laudopan or nepenthe or omnopon or oposal or pantopon* or papaveretum or pavon or tetrapon or hysingla or zohydro or lorcet or lortab or norco or vicodin or exalgo or kadian or "ms contin" or morphabond or oxaydo or roxicet or cannabis or cannabin* or cannabidiol* or marijuana* or marihuana* or endocannabinoid* or phytocannabinoid* or thc or tetrahydrocannabinol* or hash or hashish or pot) adj4 (abus* or misus* or addict* or "use disorder*")).ti. | 14844 |  |
| 40 | (COVID or COVID-19 or COVID19 or coronavirus* or corona virus* or 2019-nCoV or 19nCoV or 2019nCoV or nCoV or n-CoV or SARS-CoV-2 or SARS-CoV2 or SARSCoV-2 or SARSCoV2 or 2019-novel CoV or Sars-coronavirus2 or novel CoV).ti. | 262488 |  |
| 41 | (or/7-39) and (COVID or COVID-19 or COVID19 or coronavirus* or corona virus* or 2019-nCoV or 19nCoV or 2019nCoV or nCoV or n-CoV or SARS-CoV-2 or SARS-CoV2 or SARSCoV-2 or SARSCoV2 or 2019-novel CoV or Sars-coronavirus2 or novel CoV).ti. | 69059 | Conditions concept |
| 42 | 6 and 41 | 12180 | After + Conditions |
| 43 | *Incidence/ or *Prevalence/ or *Risk/ or *Odds Ratio/ | 5993 |  |
| 44 | (Incidence or prevalen* or probability or predict* or risk or risks or "odds ratio").ti,ab,kf. | 5447332 |  |
| 45 | ((diagno$2 or likelihood or likely) adj3 (develop* or elevated or high or higher or heightened or increase* or increasing)).ti,ab,kf. | 85600 |  |
| 46 | ((new or newly) adj3 (onset or diagnos$2 or problem$1)).ti,ab,kf. | 105135 |  |
| 47 | *Disease Progression/ or (exacerbat* or flare$1 or provoke$1 or provocation or trigger* or worsen*).ti,ab,kf. | 653197 |  |
| 48 | *Prognosis/ or (prognos?s or trajector*).ti,ab,kf. | 604857 |  |
| 49 | or/43-48 | 6368112 | IPRE Concept |
| 50 | 6 and 41 and 49 | 6881 | After + Conditions + IPRE |
| 51 | Epidemiologic Methods/ | 31606 |  |
| 52 | exp Epidemiologic Studies/ | 3019828 |  |
| 53 | Observational Studies as Topic/ | 8186 |  |
| 54 | Clinical Studies as Topic/ | 767 |  |
| 55 | (Observational Study or Clinical Study).pt. | 138383 |  |
| 56 | (observational adj3 (study or studies or design or analysis or analyses)).ti,ab,kf. | 201636 |  |
| 57 | cohort*.ti,ab,kf. | 792356 |  |
| 58 | (prospective adj3 (study or studies or design or analysis or analyses)).ti,ab,kf. | 452966 |  |
| 59 | ((follow-up or followup) adj3 (study or studies or design or analysis or analyses)).ti,ab,kf. | 94678 |  |
| 60 | ((longitudinal or long-term or longterm) adj3 (study or studies or design or analysis or analyses or data)).ti,ab,kf. | 235812 |  |
| 61 | (retrospective adj3 (study or studies or design or analysis or analyses or data or review)).ti,ab,kf. | 606375 |  |
| 62 | (case-referent adj3 (study or studies or design or analysis or analyses)).ti,ab,kf. | 633 |  |
| 63 | (population adj3 (study or studies or analysis or analyses)).ti,ab,kf. | 220933 |  |
| 64 | (descriptive adj3 (study or studies or design or analysis or analyses)).ti,ab,kf. | 100924 |  |
| 65 | ((multidimensional or multi-dimensional) adj3 (study or studies or design or analysis or analyses)).ti,ab,kf. | 4617 |  |
| 66 | ((cross-sectional or cross-sectional or crosssectional or crossectional) adj3 (study or studies or design or research or analysis or analyses or survey or findings)).ti,ab,kf. | 382982 |  |
| 67 | ((natural adj experiment) or (natural adj experiments)).ti,ab,kf. | 2965 |  |
| 68 | (quasi-experiment* or quasi-experiment*).ti,ab,kf. | 18597 |  |
| 69 | ((non-experiment$2 or nonexperiment$2) adj3 (study or studies or design or analys?s)).ti,ab,kf. | 1627 |  |
| 70 | (prevalence adj3 (study or studies or analys?s)).ti,ab,kf. | 46739 |  |
| 71 | Control groups/ | 1859 |  |
| 72 | Case-control studies/ | 323931 |  |
| 73 | ((case or group* or study or patient) adj3 (control$2 or compar* or match* or series)).ti,ab,kf. | 1529113 |  |
| 74 | Population surveillance/ | 62468 |  |
| 75 | (surveillance adj3 (continuous or continual or continued or data or ongoing or population or research or study)).ti,ab,kf. | 32513 |  |
| 76 | or/51-75 | 5029016 |  |
| 77 | 50 and 76 | 3951 | Study filter applied |
| 78 | (Case Reports.pt. or (case report? or case study or case studies).ti.) not (review* or trial*).ti,ab,kf,hw. | 2095564 |  |
| 79 | comment/ or editorial/ or (comment or editorial or news or newspaper article).pt. | 1621522 |  |
| 80 | (exp Animals/ or Models, Animal/ or Disease Models, Animal/) not Humans/ | 5053146 |  |
| 81 | ((animal or animals or canine* or dog or dogs or feline or hamster* or lamb or lambs or mice or monkey or monkeys or mouse or murine or pig or pigs or piglet* or porcine or primate* or rabbit* or rats or rat or rodent* or sheep* or veterinar*) not (human* or patient*)).mp. | 4915740 |  |
| 82 | 77 not (or/78-81) | 3837 |  |
| 83 | limit 82 to yr="2020 -Current" | 3834 |  |
| 84 | remove duplicates from 83 | 3750 |  |

##### EMBASE Search strategy – After Covid: Health conditions development/exacerbation

**After Concept**

1 - draft - Covid chronic - After concept_MEDLINE_EMBASE

**Conditions Concept**

1 - draft - Covid chronic - chronic disease_PHAC KW3_ EMBASE

**Incidence/Prev/Risk Concept**

1 - draft - Covid chronic - chronic disease_IPRE3_ EMBASE

**Study Design Filter**

1 - draft - Covid chronic - study design filter_EMBASE

**Fluff Filter**

1 - draft - Covid chronic - Animal and fluff filter_EMBASE

Database(s): Embase 1974 to 2022 October 03
Search Strategy: After Covid_Health Conditions_EMBASE_4Oct2022

Date Run: 4Oct2022

| **#** | **Searches** | **Results** | **Annotations** |
| --- | --- | --- | --- |
| 1 | ("after covid" or post-covid or postcovid or post-coronavir* or post-coronavir*).ti,ab. | 10971 |  |
| 2 | ((post-acute or postacute or "after acute" or "after discharge" or "after hospital discharge" or sequela* or post-infect* or post-viral or postviral or post-discharg* or postdischarge or survivor$1) adj5 (COVID or COVID-19 or COVID19 or coronavirus* or corona virus* or 2019-nCoV or 19nCoV or 2019nCoV or nCoV or n-CoV or SARS-CoV-2 or SARS-CoV2 or SARSCoV-2 or SARSCoV2 or 2019-novel CoV or Sars-coronavirus2 or novel CoV)).ti,ab. | 4650 |  |
| 3 | (("more than" or longer or "at least" or "after" or follow* or upward* or later* or prospectiv* or retrospectiv* or review* or longitud* or post or beyond) adj5 (day? or week? or wk or wks or month? or mo or mos or year? or yr or yrs)).ti,ab. | 4042093 |  |
| 4 | (COVID or COVID-19 or COVID19 or coronavirus* or corona virus* or 2019-nCoV or 19nCoV or 2019nCoV or nCoV or n-CoV or SARS-CoV-2 or SARS-CoV2 or SARSCoV-2 or SARSCoV2 or 2019-novel CoV or Sars-coronavirus2 or novel CoV).ti. | 280853 |  |
| 5 | 3 and 4 | 32735 |  |
| 6 | 1 or 2 or 5 | 41972 | After concept |
| 7 | (Chronic adj3 (condition$1 or diagnos$2 or disease$1 or disorder* or failure or illness*)).ti,ab,kw. | 678865 |  |
| 8 | ((longterm or long-term) adj3 (consequence* or damage or effect$1 or impact* or outcome* or result* or sequela*)).ti,ab,kw. | 363475 |  |
| 9 | ("cardiovascular stroke*" or "heart attack*" or "heart infarct*" or "myocardial infarct*").ti,ab. | 316429 |  |
| 10 | "myocardial revasculari?ation".ti,ab. | 6336 |  |
| 11 | (((cardio* or cardia* or heart? or myocardi* or vascula* or atria* or ventric* or coronary) and (disease* or disorder* or condition* or syndrome* or illness* or abnormal* or dysfunction* or disturb* or event* or adverse* or inflam*)) or nstemi or stemi or cvd or fibrillation or paroxysmal dyspnea or arrhythmia* or dysrhythmia* or bradycard* or brugada syndrome* or commotio cordis or long qt syndrome or parasystole or pre-excitation syndrome* or tachycardia* or conduction disturbance*).ti,ab. | 2414948 |  |
| 12 | ("cerebrovascular accident*" or stroke or strokes).ti,ab. | 476487 |  |
| 13 | (diabet* or "t1dm" or "t1d" or "t2dm" or "t2d" or "niddm" or insulin resistan* or prediabet* or dysglyc?emi* or hyperglyc?emi* or hypoglyc?emi* or hyper glyc?emi* or hypo glyc?emi* or "hyperosmolar state*" or ketoacidosis or (impaired fasting adj2 (glucose or sugar)) or ((glucose or carbohydrate*) adj3 (toler* or intoler*))).ti,ab. | 1245962 |  |
| 14 | ((chronic obstructive adj4 (pulmonary or airway? or lung?)) or chronic airflow obstruction* or copd or coad or aecopd or obstructive pulmonary or asthma* or emphysema* or (chronic adj3 (bronchitis or bronchus infection* or bronchi infection* or bronchus inflammation or bronchi inflammation))).ti,ab. | 398637 |  |
| 15 | (osteopor* or arthrit* or antisynthetase syndrome or behcet* disease or blau syndrom* or monarthrit* or monoarthrit* or coxitis* or sacroiliitis or osteoarthrit* or arthrosis* or spondylosis* or barre criecou or barre lieou or brachialgia paraesthetica nocturna or brachialgia paresthetica nocturna or neri barre or osteophytosis or polyspondylitis or spondylarthrosis or spondyloarthrosis or papa syndrome or papga syndrome or pigmented villonodular synovitis or "pvns" or reiter syndrome* or arthragra* or gout* or cheiragra* or chiragra* or urate inflammation* or pseudogout* or (crystal* adj3 arthropath*)).ti,ab. | 552499 |  |
| 16 | (epilep* or seizure* or panayiotopoulos syndrome or rasmussen syndrome or alpers disease or hypsarrhythmia or infantie spasm* or lennox gastaut syndrome* or merrf syndrome* or myoclonus* or nodding syndrome* or lipofuscinosis or (sclerosis adj3 (multiple* or disseminate* or insular*)) or chariot disease* or parkinsonism* or hemiparkinsonism* or (parkinson* adj3 (syndrom* or disorder* or disease*)) or paralysis agitans or amnesia* or delirium or alzheimer* or aphasi* or diffuse neurofibrillary tangle* or lobar degeneration* or acquired dyslexia or alexia or cadasil or cada sil or cerebral autosomal dominant arteriopathy or lewy body disease* or (pick* adj1 (complex* or disease* or disorder* or syndrome*)) or huntington* or juvenile chorea or chorea major or kluver bucy or (mental* adj3 deteriorat*) or pseudodementia* or rett syndrome* or senile* or tauopath* or ((neuro* or neural* or cognitive* or cognition* or consciousness or sleep* or wake*) adj3 (impair* or side effect* or function* or disfunction* or dysfunction* or disorder* or degenerat* or disease*))).ti,ab. | 1549063 |  |
| 17 | (((chronic kidney or chronic renal) adj2 (disease? or insufficienc* or failur?)) or CKD or chronic nephropath*).ti,ab. | 161815 |  |
| 18 | (((cancer* or tumor? or tumour?) adj3 (induct* or form* or genesis or caus*)) or cancerogen* or neoplasmogenesis or oncogenesis or oncogenicit* or tumorigenicit* or tumorigenesis or tumourgenicit* or tumourigenesis or carcinogeni* or (cell adj3 proliferat*) or clonal expan* or (foci adj3 alter*) or (reactivat* adj4 dorman*) or adamantinoma* or adenoacanthoma* or adenocanthoma* or adenocarcinoma* or adenomatos* or adenomyoepithelioma* or adenomyoma* or adenosarcoma* or aesthesioneuroblastoma* or ameloblastoma* or androblastoma* or angioblastoma* or angioendothelioma* or angiofibrosarcoma* or angiosarcoma* or argentaffinoma* or arrhenoblastoma* or astroblastoma* or astrocytoma* or astroglioma* or baltoma or basiloma* or basalioma* or blastoma? or buschke-lowenstein or cancer* or carcinogen* or carcinoid? or carcinoma* or carcinosarcoma* or chloroma* or cholangiocarcinoma* or cholangiohepatoma* or cholangiosarcoma* or chondroblastoma* or chondrosarcoma* or chorioadenoma* or chorioangioma or choriocarcinoma* or chorioepithelioma* or chorionepithelioma* or comedocarcinoma* or cystadenocarcinoma* or cystosarcoma* or dermatofibrosarcoma* or dictyoma* or dysgerminoma* or ectomesenchymoma* or ependymoblastoma* or erythroleukaemia* or erythroleukemia* or erythroplakia* or esthesioneuroblastoma* or esthesioneuroepithelioma* or fibroadenosarcoma* or fibrochondrosarcoma* or fibroepithelioma* or fibroliposarcoma* or fibrosarcoma* or fibroxanthoma* or fibroxanthosarcoma* or ganglioblastoma* or ganglioneuroblastoma* or glioblastoma* or gliomatos* or gliosarcoma* or haemangioblastoma* or haemangiopericytoma* or haemangiosarcoma* or hemangioblastoma* or hemangioendothelioma* or hemangioendotheliosarcoma* or hemangiopericytoma* or hemangiosarcoma* or hepatoblastoma* or hepatocarcinoma* or hepatocholangiocarcinoma* or hepatoma? or hodgkin* or hypernephroma* or immunocytoma* or keratoacanthoma* or leiomyosarcoma* or leukaemia* or leukemia* or liposarcoma* or lymphoma? or lymphangiosarcoma* or lymphoepithelioma* or macroglobulinemia* or malignan* or maltoma* or medulloblastoma* or medullomyoblastoma* or melanoameloblastoma* or melanoma* or meningioblastoma* or mesenchymoma* or mesonephroma* or mesothelioma* or metasta* or microglioma* or myelodysplas* or myeloma* or myelomatos* or myeloproliferat* or myosarcoma* or myxoliposarcoma* or nephroblastoma* or neuroblastoma* or neurofibrosarcoma* or nonhodgkin* or nonseminoma* or oligodendroglioma* or oncocytoma* or oncogen* or oncolog* or orchioblastoma* or osteoblastoma* or osteochondrosarcoma* or osteofibrosarcoma* or osteosarcoma* or pancreatoblastoma* or paraganglioma* or paraneoplastic or pinealblastoma* or pinealoblastoma* or pineoblastoma* or pneumoblastoma* or polyembryoma* or polyhistioma* or porocarcinoma* or precancer* or preleukaemia* or preleukemia* or premalignant* or preneoplastic* or reticuloendothelioma* or retinoblastoma* or rhabdomyosarcomas* or rhabdosarcoma* or sarcoma* or seminoma* or somatostatinoma* or teratocarcinoma* or waldenstrom* or xanthosarcoma*).ti,ab. | 5283528 |  |
| 19 | (anxiety* or depression* or schizo* or psychotic* or psychosis or psychoses or amnesia* or delirium* or gender dysphoria* or paraphilia* or tourette* or hallucination* or paranoid or paranoia or paraphrenia or sexual dysfunction* or hysteria or phobia? or agoraphobia or neurasthenia or hypochondriasis or hypochondria or hypochondriac or adjustment reaction*).ti,ab. | 1014019 |  |
| 20 | ((manic or mania or depress*) adj3 episode*).ti,ab. | 21539 |  |
| 21 | ((bipolar* or personality or mood* or conduct or impulse or dissociative or eating or neurotic or obsessive or compulsive or panic or phobic or neurocognitiv* or trauma* or posttrauma* or stress* or neurocognitive* or amnestic* or behaviour* or behavior* or neurobehav* or delusion* or manic or mania or depress* or conversion or somatoform or gender identity or attention-deficit or hyperactivity or conduct or emotion* or tic or tics or sleep or sexual identity or dysthymic or alcohol-induced or drug-induced or social functioning) adj3 disorder*).ti,ab. | 404682 |  |
| 22 | ((mental* or psychiatric* or psychological* or neuropsych*) adj3 (disorder* or ill or illness* or diagnosis or diagnoses)).ti,ab. | 241550 |  |
| 23 | ((intellectual* or developmental* or neurodevelopmental or learning) adj3 (disabilit* or disorder* or syndrome*)).ti,ab. | 90894 |  |
| 24 | (alcoholism* or ((glue or gas) adj3 (sniffing or huffing)) or ((nicotine or tobacco) adj3 (dependen* or abus* or misus* or addict* or overdos* or over dos* or "use disorder*"))).ti,ab. | 57704 |  |
| 25 | ((drug? or substance? or multidrug? or polydrug? or polysubstance? or stimulant? or alcohol or sedative* or hypnotic* or anxiolytic* or psychoactive*) adj4 (abus* or misus* or addict* or "use disorder*")).ti,ab. | 172706 |  |
| 26 | ((adderall or angel dust or amphetamine? or ayahuasca or barbiturate* or benzodiazepine* or buprenorphine or crack or cocaine or codeine or demerol or dextroamphetamine or dextromethamphetamine or diamorphine or dihydromorphine or dilaudid or ecstasy or ethylmorphine or fentanyl or hallucinogen* or heroin or hydrocodeine or hydromorphone or hyrdocodone or ketamine or kratom or lsd or lysergic acid diethylamide or magic mint or mdma or meperidine or mescaline or methadone or meth or methamphetamine or methylenedioxyamphetamine or methylenedioxymethamphetamine or methylphenidate or methylphenidate or midomafetamine or molly or naltrexone or pcp or pethidine or peyote or phencyclidine or psilocybin or ritalin or salvia or suboxone or tramadol or ghb or gamma?hydroxybutyrate or inhalent? or inhalant? or narcotic? or opioid* or opiate* or opium or opon or dextropropoxyphene or anileridine or alphaprodine or diamorphine or levorphanol or sufentanil or bromadol or bdpc or etorphine or dihydroetorphine or carfentanil or heroin or alfentanil or remifentanil or etorphine or buprenorphine or anorfin or buprenex or buprex or buprine or butrans or norphin or pentorel or prefin or probuphine or subutex or temgesic or transtec or bupe or bute or suboxone or zubsolv or subby or sobos or codein? or codipertussin or codyl or methylmorfine or methylmorphine or pentuss or "tylenol 2" or "tylenol 3" or "tylenol 4" or cody or fentanyl or abstral or duragesic or durogesic or fentamyl or fentanil or fetnanyl or instanyl or ionsys or lazanda or leptanal or onsolis or pecfent or phentanyl or rapinyl or recuvyra or subsys or tanyl or transfenta or onsolis or bekadid or dico or dicodid or dihydrocodeinone or dihydrocodone or hydrocodone* or hydrocodonum or tussionex or vicoprofen or vike or hydromorphone or dilaudid or dillies or meperidine or pethidine or algil or alodan or centralgin or centralgine or cluyer or demero or demerol or dispadol or dolanquifa or dolantal or dolantin or dolantina or dolantine or dolargan or dolcontral or dolenal or dolestin or dolestine or dolin or dolocontral or doloneurin or doloneurotrat or dolor or dolosa or dolosal or dolosan or dolsin or dolvanol or endolate or isonipecaine or lidol or lydol or mefedina or mepadin or meperdol or meperiden or meperidin or meperidol or mephedine or mepiridine or mialgin or neomochin or opistan or pantalgin or petadin or petantin or petantina or pethanol or pethedine or pethidin or petidin or phetidine or piridosal or sauteralgyl or simesalgina or supplosal or synlaudine or demmies or methadone or adanon or algidon or algolysin or algoxale or althose or amidon or amidona or amidone or amidosan or anadon or biodone or butalgin or deamin or depridol or diaminon or dianone or dolafin or dolamid or dolesone or dolmed or dolophine or dorex or dorexol or eptadone or fenadon or gobbidona or heptadon or heptanon or ketalgin or mecodin or mepecton or mephenon or metadol or metadon or metasedin or methaddict or methadon or methadose or "methaforte mix" or miadone or moheptan or pallidone or phenadon or physepton or physeptone or polamidon or polamivet or polamivit or sinalgin or symoron or westadone or meth or morphine or anpec or duromorph or epimorph or miro or morfin or morfine or morphia or morphin or morphinium or morphium or opso or skenan or doloral or statex or "m.o.s." or morph or "red rockets" or oxycodone or bionine or bionone or bolodorm or broncodal or bucodal or cafacodal or cardanon or codenon or dihydrohydroxycodeinone or dihydrohydroxydodeinone or dihydrone or dinarkon or endone or eubine or eucodal or eucodale or eucodalum or eudin or eukdin or eukodal or eumorphal or eurodamine or eutagen or hydrocodal or hydroxycodeinoma or ludonal or "m-oxy" or medicodal or narcobasina or narcobasine or narcosin or nargenol or narodal or nucodan or opton or ossicodone or oxanest or oxecta or oxicone or oxicontin or oxiconum or oxikon or "oxy ir" or oxycod or oxycodeinonhydrochloride or oxycodyl or oxycone or oxycontin or oxydose or oxyfast or oxygesic or oxyir or oxykon or oxynorm or pancodine or pavinal or percolone or pronarcin or remoxy or roxicodone or roxycodone or sinthiodal or stupenal or supeudol or tebodal or tekodin or thecodin or oxyneo or percocet or oxycocet or percodan or oxy or percs or pentazocine or dolapent or fortal or fortalgesic or fortral or fortraline or fortwin or liticon or peltazon or pentacozine or pentafen or pentagin or pentalgina or pentazocin or pentozocine or perutagin or sosegon or sosigon or talioin or talwin or tapentadol or nucynta or palexia or yantil or nucynta or tramadol or adamon or amanda or analab or analdol or andalpha or bellatram or biodalgic or calmador or calmol or contramal or dolana or dolika or dolmal or dolotral or dolzam or dromadol or eufindol or exopen or katrasic or mabron or melanate or mosepan or newdorphin or nobligan or nonalges or omnidol or pengesic or penimadol or prontofort or radol or rofy or ryzolt or sefmal or sensitram or takadol or tamolan or tandol or tarol or topalgic or trabar or trabilan or trabilin or tradol or tradolan or tradonal or tralic or tramada or tramadex or tramagetic or tramagit or tramahexal or tramake or tramal or tramazac or tramed or tramol or tramundin or trasedal or trasik or "trd-contin" or trexol or tridol or trodon or trondon or ultram or unitral or urgendol or zamadol or zamudol or zodol or zumatran or zydol or tramacet or tridural or durela or "chill pill?" or oxymorphone or opana or algopan or quarelin or actiq or fentora or biopan or cofapon or laudopan or nepenthe or omnopon or oposal or pantopon* or papaveretum or pavon or tetrapon or hysingla or zohydro or lorcet or lortab or norco or vicodin or exalgo or kadian or "ms contin" or morphabond or oxaydo or roxicet or cannabis or cannabin* or cannabidiol* or marijuana* or marihuana* or endocannabinoid* or phytocannabinoid* or thc or tetrahydrocannabinol* or hash or hashish or pot) adj4 (abus* or misus* or addict* or "use disorder*")).ti. | 17796 |  |
| 27 | (or/7-26) and (COVID or COVID-19 or COVID19 or coronavirus* or corona virus* or 2019-nCoV or 19nCoV or 2019nCoV or nCoV or n-CoV or SARS-CoV-2 or SARS-CoV2 or SARSCoV-2 or SARSCoV2 or 2019-novel CoV or Sars-coronavirus2 or novel CoV).ti. | 71793 | Conditions concept |
| 28 | 6 and 27 | 16235 | After + Conditions |
| 29 | (Incidence or prevalen* or probability or predict* or risk or risks or "odds ratio").ti,ab,kw. | 7450177 |  |
| 30 | ((diagno$2 or likelihood or likely) adj3 (develop* or elevated or high or higher or heightened or increase* or increasing)).ti,ab,kw. | 122050 |  |
| 31 | ((new or newly) adj3 (onset or diagnos$2 or problem$1)).ti,ab,kw. | 186710 |  |
| 32 | (exacerbat* or flare$1 or provoke$1 or provocation or trigger* or worsen*).ti,ab,kf. | 917818 |  |
| 33 | (prognos?s or trajector*).ti,ab,kw. | 867733 |  |
| 34 | or/29-33 | 8695941 | IPRE Concept |
| 35 | 6 and 27 and 34 | 9876 | After + Conditions + IPRE |
| 36 | observational study/ | 290223 |  |
| 37 | cohort analysis/ | 902797 |  |
| 38 | longitudinal study/ | 179148 |  |
| 39 | follow up/ | 1903266 |  |
| 40 | retrospective study/ | 1316750 |  |
| 41 | exp case control study/ | 211745 |  |
| 42 | cross-sectional study/ | 508502 |  |
| 43 | quasi experimental study/ | 10033 |  |
| 44 | prospective study/ | 799288 |  |
| 45 | (observational adj3 (study or studies or design or analysis or analyses)).ti,ab,kf. | 312637 |  |
| 46 | cohort*.ti,ab,kf. | 1333150 |  |
| 47 | (prospective adj3 (study or studies or design or analysis or analyses)).ti,ab,kf. | 675354 |  |
| 48 | ((follow up or followup) adj3 (study or studies or design or analysis or analyses)).ti,ab,kf. | 138278 |  |
| 49 | ((longitudinal or long-term or longterm) adj3 (study or studies or design or analysis or analyses or data)).ti,ab,kf. | 324572 |  |
| 50 | (retrospective adj3 (study or studies or design or analysis or analyses or data or review)).ti,ab,kf. | 1008244 |  |
| 51 | (case-referent adj3 (study or studies or design or analysis or analyses)).ti,ab,kf. | 694 |  |
| 52 | (population adj3 (study or studies or analysis or analyses)).ti,ab,kf. | 328917 |  |
| 53 | (descriptive adj3 (study or studies or design or analysis or analyses)).ti,ab,kf. | 152114 |  |
| 54 | ((multidimensional or (multi adj dimensional)) adj3 (study or studies or design or analysis or analyses)).ti,ab,kf. | 5367 |  |
| 55 | ((cross-sectional or cross-sectional or crosssectional or crossectional) adj3 (study or studies or design or research or analysis or analyses or survey or findings)).ti,ab,kf. | 502411 |  |
| 56 | ((natural adj experiment) or (natural adj experiments)).ti,ab,kf. | 3200 |  |
| 57 | (quasi-experiment* or quasi-experiment*).ti,ab,kf. | 23009 |  |
| 58 | ((non-experiment$2 or nonexperiment$2) adj3 (study or studies or design or analys?s)).ti,ab,kf. | 2191 |  |
| 59 | (prevalence adj3 (study or studies or analys?s)).ti,ab,kf. | 67060 |  |
| 60 | control group/ | 110512 |  |
| 61 | case control study/ | 193533 |  |
| 62 | ((case or group* or study or patient) adj3 (control$2 or compar* or match* or series)).ti,ab,kf. | 2187788 |  |
| 63 | population surveillance/ | 137 |  |
| 64 | (surveillance adj3 (continuous or continual or continued or data or ongoing or population or research or study)).ti,ab,kf. | 43080 |  |
| 65 | or/36-64 | 7176339 |  |
| 66 | 35 and 65 | 6609 | Study design applied |
| 67 | (Case Reports.pt. or (case report? or case study or case studies).ti.) not (review* or trial*).ti,ab,kf,hw. | 326140 |  |
| 68 | comment/ or editorial/ or (comment or editorial).pt. | 781556 |  |
| 69 | (conference abstract* or conference paper*).af. not (conference abstract* or conference paper*).ab,ga. | 5336419 |  |
| 70 | conference proceeding*.af. not conference proceeding*.ab,ga. | 39394 |  |
| 71 | (exp animal/ or exp animal model/) not human/ | 5262807 |  |
| 72 | ((animal or animals or canine* or dog or dogs or feline or hamster* or lamb or lambs or mice or monkey or monkeys or mouse or murine or pig or pigs or piglet* or porcine or primate* or rabbit* or rats or rat or rodent* or sheep* or veterinar*) not (human* or patient*)).mp. | 4822166 |  |
| 73 | 66 not (or/67-72) | 3867 |  |
| 74 | limit 73 to yr="2020 -Current" | 3865 |  |
| 75 | remove duplicates from 74 | 3812 |  |

#### Appendix 3. Excluded studies lists, by reasons for exclusion.

##### Excluded due to lack of age-stratification

Al-Aly Z, Bowe B, Xie Y. Long COVID after breakthrough SARS-CoV-2 infection. *Nature medicine* 2022;28(7):1461-67. doi: https://dx.doi.org/10.1038/s41591-022-01840-0

Al-Aly Z, Xie Y, Bowe B. High-dimensional characterization of post-acute sequelae of COVID-19. *Nature* 2021;594(7862):259-64. doi: https://dx.doi.org/10.1038/s41586-021-03553-9

Bhaskaran K, Rentsch CT, Hickman G, et al. Overall and cause-specific hospitalisation and death after COVID-19 hospitalisation in England: A cohort study using linked primary care, secondary care, and death registration data in the OpenSAFELY platform. *PLoS medicine* 2022;19(1):e1003871. doi: https://dx.doi.org/10.1371/journal.pmed.1003871

Birabaharan M, Kaelber DC, Pettus JH, et al. Risk of new-onset type 2 diabetes in 600 055 people after COVID-19: A cohort study. *Diabetes, obesity & metabolism* 2022;24(6):1176-79. doi: https://dx.doi.org/10.1111/dom.14659

Bsteh G, Assar H, Gradl C, et al. Long-term outcome after COVID-19 infection in multiple sclerosis: a nation-wide multicenter matched-control study. *European journal of neurology* 2022 doi: https://dx.doi.org/10.1111/ene.15477

Cecchetti G, Agosta F, Canu E, et al. Cognitive, EEG, and MRI features of COVID-19 survivors: a 10-month study. *Journal of Neurology* 2022;269(7):3400-12. doi: https://dx.doi.org/10.1007/s00415-022-11047-5

Cian V, De Laurenzis A, Siri C, et al. Cognitive and Neuropsychiatric Features of COVID-19 Patients After Hospital Dismission: An Italian Sample. *Frontiers in psychology* 2022;13:908363. doi: https://dx.doi.org/10.3389/fpsyg.2022.908363

Clift AK, Ranger TA, Patone M, et al. Neuropsychiatric Ramifications of Severe COVID-19 and Other Severe Acute Respiratory Infections. *JAMA psychiatry* 2022;79(7):690-98. doi: https://dx.doi.org/10.1001/jamapsychiatry.2022.1067

Coleman B, Casiraghi E, Blau H, et al. Increased risk of psychiatric sequelae of COVID-19 is highest early in the clinical course. *medRxiv : the preprint server for health sciences* 2021 doi: https://dx.doi.org/10.1101/2021.11.30.21267071

Di Maio S, Lamina C, Coassin S, et al. Lipoprotein(a) and SARS-CoV-2 infections: Susceptibility to infections, ischemic heart disease and thromboembolic events. *Journal of internal medicine* 2022;291(1):101-07. doi: https://dx.doi.org/10.1111/joim.13338

Hentschel CB, Abramoff BA, Dillingham TR, et al. Race, ethnicity, and utilization of outpatient rehabilitation for treatment of post COVID-19 condition. *PM & R : the journal of injury, function, and rehabilitation* 2022 doi: https://dx.doi.org/10.1002/pmrj.12869

Ingul CB, Grimsmo J, Mecinaj A, et al. Cardiac Dysfunction and Arrhythmias 3 Months After Hospitalization for COVID-19. *Journal of the American Heart Association* 2022;11(3):e023473. doi: https://dx.doi.org/10.1161/JAHA.121.023473

Kennedy J, Parker M, Seaborne M, et al. Health care use attributable to COVID-19: A propensity matched national electronic health records cohort study of 249,390 people in Wales, UK. *medRxiv* 2022 doi: https://dx.doi.org/10.1101/2022.04.21.22274152

Kerchberger VE, Peterson JF, Wei W-Q. Scanning the medical phenome to identify new diagnoses after recovery from COVID-19 in a US cohort. *Journal of the American Medical Informatics Association : JAMIA* 2022 doi: https://dx.doi.org/10.1093/jamia/ocac159

Kim N, Kim J, Yang BR, et al. Associations of unspecified pain, idiopathic pain and COVID-19 in South Korea: a nationwide cohort study. *The Korean journal of pain* 2022;35(4):458-67. doi: https://dx.doi.org/10.3344/kjp.2022.35.4.458

Knight R, Walker V, Ip S, et al. Association of COVID-19 With Major Arterial and Venous Thrombotic Diseases: A Population-Wide Cohort Study of 48 Million Adults in England and Wales. *Circulation* 2022;146(12):892-906. doi: https://dx.doi.org/10.1161/CIRCULATIONAHA.122.060785

Lal BK, Prasad NK, Englum BR, et al. Periprocedural complications in patients with SARS-CoV-2 infection compared to those without infection: A nationwide propensity-matched analysis. *American journal of surgery* 2021;222(2):431-37. doi: https://dx.doi.org/10.1016/j.amjsurg.2020.12.024

Lip GYH, Genaidy A, Tran G, et al. Effects of multimorbidity on incident COVID-19 events and its interplay with COVID-19 event status on subsequent incident myocardial infarction (MI). *European journal of clinical investigation* 2022;52(5):e13760. doi: https://dx.doi.org/10.1111/eci.13760

Moga TD, Nistor-Cseppento CD, Bungau SG, et al. The Effects of the 'Catabolic Crisis' on Patients' Prolonged Immobility after COVID-19 Infection. *Medicina (Kaunas, Lithuania)* 2022;58(6) doi: https://dx.doi.org/10.3390/medicina58060828

Mohanka MR, Mahan LD, Joerns J, et al. Clinical characteristics, management practices, and outcomes among lung transplant patients with COVID-19. *The Journal of heart and lung transplantation : the official publication of the International Society for Heart Transplantation* 2021;40(9):936-47. doi: https://dx.doi.org/10.1016/j.healun.2021.05.003

Murata F, Maeda M, Ishiguro C, et al. Acute and delayed psychiatric sequelae among patients hospitalised with COVID-19: a cohort study using LIFE study data. *General psychiatry* 2022;35(3):e100802. doi: https://dx.doi.org/10.1136/gpsych-2022-100802

Nersesjan V, Fonsmark L, Christensen RHB, et al. Neuropsychiatric and Cognitive Outcomes in Patients 6 Months After COVID-19 Requiring Hospitalization Compared With Matched Control Patients Hospitalized for Non-COVID-19 Illness. *JAMA psychiatry* 2022;79(5):486-97. doi: https://dx.doi.org/10.1001/jamapsychiatry.2022.0284

Nersesjan V, Amiri M, Christensen HK, et al. Thirty-Day Mortality and Morbidity in COVID-19 Positive vs. COVID-19 Negative Individuals and vs. Individuals Tested for Influenza A/B: A Population-Based Study. *Frontiers in medicine* 2020;7:598272. doi: https://dx.doi.org/10.3389/fmed.2020.598272

Oh TK, Park HY, Song IA. Risk of psychological sequelae among coronavirus disease-2019 survivors: A nationwide cohort study in South Korea. *Depression and Anxiety* 2021;38(2):247-54. doi: https://dx.doi.org/10.1002/da.23124

Patel N, Dahman B, Bajaj JS. Development of New Mental and Physical Health Sequelae among US Veterans after COVID-19. *Journal of clinical medicine* 2022;11(12) doi: https://dx.doi.org/10.3390/jcm11123390

Prasad NK, Lake R, Englum BR, et al. Increased complications in patients who test COVID-19 positive after elective surgery and implications for pre and postoperative screening. *American journal of surgery* 2022;223(2):380-87. doi: https://dx.doi.org/10.1016/j.amjsurg.2021.04.005

Priyadarshni S, Westra J, Kuo Y-F, et al. COVID-19 Infection and Incidence of Myocarditis: A Multi-Site Population-Based Propensity Score-Matched Analysis. *Cureus* 2022;14(2):e21879. doi: https://dx.doi.org/10.7759/cureus.21879

Rathmann W, Kuss O, Kostev K. Incidence of newly diagnosed diabetes after Covid-19. *Diabetologia* 2022;65(6):949-54. doi: https://dx.doi.org/10.1007/s00125-022-05670-0

Rivera-Izquierdo M, Lainez-Ramos-Bossini AJ, de Alba IG-F, et al. Long COVID 12 months after discharge: persistent symptoms in patients hospitalised due to COVID-19 and patients hospitalised due to other causes-a multicentre cohort study. *BMC medicine* 2022;20(1):92. doi: https://dx.doi.org/10.1186/s12916-022-02292-6

Salah HM, Fudim M, O'Neil ST, et al. Post-recovery COVID-19 and incident heart failure in the National COVID Cohort Collaborative (N3C) study. *Nature communications* 2022;13(1):4117. doi: https://dx.doi.org/10.1038/s41467-022-31834-y

Shabaka A, Gruss E, Landaluce-Triska E, et al. Late thrombotic complications after SARS-CoV-2 infection in hemodialysis patients. *Hemodialysis international International Symposium on Home Hemodialysis* 2021;25(4):507-14. doi: https://dx.doi.org/10.1111/hdi.12935

Sharma A, Misra-Hebert AD, Mariam A, et al. Impacts of COVID-19 on Glycemia and Risk of Diabetic Ketoacidosis. *Diabetes* 2022 doi: https://dx.doi.org/10.2337/db22-0264

Sorensen AIV, Spiliopoulos L, Bager P, et al. Post-acute symptoms, new onset diagnoses and health problems 6 to 12 months after SARS-CoV-2 infection: a nationwide questionnaire study in the adult Danish population. *medRxiv* 2022 doi: https://dx.doi.org/10.1101/2022.02.27.22271328

Spencer-Segal JL, Smith CA, Slavin A, et al. Mental health outcomes after hospitalization with or without COVID-19. *General hospital psychiatry* 2021;72:152.

Strohbehn IA, Zhao S, Seethapathy H, et al. Acute Kidney Injury Incidence, Recovery, and Long-term Kidney Outcomes Among Hospitalized Patients With COVID-19 and Influenza. *Kidney international reports* 2021;6(10):2565-74. doi: https://dx.doi.org/10.1016/j.ekir.2021.07.008

Sun S, Annadi RR, Chaudhri I, et al. Short- and Long-Term Recovery after Moderate/Severe AKI in Patients with and without COVID-19. *Kidney360* 2022;3(2):242-57. doi: https://dx.doi.org/10.34067/KID.0005342021

Taquet M, Dercon Q, Luciano S, et al. Incidence, co-occurrence, and evolution of long-COVID features: A 6-month retrospective cohort study of 273,618 survivors of COVID-19. *PLoS medicine* 2021;18(9):e1003773. doi: https://dx.doi.org/10.1371/journal.pmed.1003773

Taquet M, Geddes JR, Husain M, et al. 6-month neurological and psychiatric outcomes in 236 379 survivors of COVID-19: a retrospective cohort study using electronic health records. *The lancet Psychiatry* 2021;8(5):416-27. doi: https://dx.doi.org/10.1016/S2215-0366(21)00084-5

Tereshchenko LG, Bishop A, Fisher-Campbell N, et al. Risk of Cardiovascular Events After COVID-19. *The American journal of cardiology* 2022;179:102-09. doi: https://dx.doi.org/10.1016/j.amjcard.2022.06.023

Tirozzi A, Santonastaso F, de Gaetano G, et al. Does COVID-19 increase the risk of neuropsychiatric sequelae? Evidence from a mendelian randomization approach. *World journal of psychiatry* 2022;12(3):536-40. doi: https://dx.doi.org/10.5498/wjp.v12.i3.536

Tsai S, Nguyen H, Ebrahimi R, et al. COVID-19 associated mortality and cardiovascular disease outcomes among US women veterans. *Scientific reports* 2021;11(1):8497. doi: https://dx.doi.org/10.1038/s41598-021-88111-z

Wander PL, Lowy E, Beste LA, et al. The Incidence of Diabetes Among 2,777,768 Veterans With and Without Recent SARS-CoV-2 Infection. *Diabetes care* 2022;45(4):782-88. doi: https://dx.doi.org/10.2337/dc21-1686

Ward A, Sarraju A, Lee D, et al. COVID-19 is associated with higher risk of venous thrombosis, but not arterial thrombosis, compared with influenza: Insights from a large US cohort. *PloS one* 2022;17(1):e0261786. doi: https://dx.doi.org/10.1371/journal.pone.0261786

Xiang Y, Zhang R, Qiu J, et al. Association of COVID-19 with risks of hospitalization and mortality from other disorders post-infection: A study of the UK Biobank. *medRxiv* 2022 doi: https://dx.doi.org/10.1101/2022.03.23.22272811

Xie Y, Xu E, Al-Aly Z. Risks of mental health outcomes in people with covid-19: cohort study. *BMJ (Clinical research ed)* 2022;376:e068993. doi: https://dx.doi.org/10.1136/bmj-2021-068993

Zhang HG, Dagliati A, Shakeri Hossein Abad Z, et al. International electronic health record-derived post-acute sequelae profiles of COVID-19 patients. *NPJ digital medicine* 2022;5(1):81. doi: https://dx.doi.org/10.1038/s41746-022-00623-8

##### Excluded for insufficient adjustment in analysis

Barrett CE, Koyama AK, Alvarez P, et al. Risk for Newly Diagnosed Diabetes >30 Days After SARS-CoV-2 Infection Among Persons Aged <18 Years - United States, March 1, 2020-June 28, 2021. *MMWR Morbidity and mortality weekly report* 2022;71(2):59-65. doi: https://dx.doi.org/10.15585/mmwr.mm7102e2

Bull-Otterson L, Baca S, Saydah S, et al. Post–COVID Conditions Among Adult COVID-19 Survivors Aged 18–64 and≥ 65 Years—United States, March 2020–November 2021. *Morbidity and Mortality Weekly Report* 2022;71(21):713.

Estiri H, Strasser ZH, Brat GA, et al. Evolving phenotypes of non-hospitalized patients that indicate long COVID. *BMC medicine* 2021;19(1):249. doi: https://dx.doi.org/10.1186/s12916-021-02115-0

Graham EL, Clark JR, Orban ZS, et al. Persistent neurologic symptoms and cognitive dysfunction in non‐hospitalized Covid‐19 “long haulers”. *Annals of clinical and translational neurology* 2021;8(5):1073-85.

Hernandez-Romieu AC, Carton TW, Saydah S, et al. Prevalence of Select New Symptoms and Conditions Among Persons Aged Younger Than 20 Years and 20 Years or Older at 31 to 150 Days After Testing Positive or Negative for SARS-CoV-2. *JAMA network open* 2022;5(2):e2147053. doi: https://dx.doi.org/10.1001/jamanetworkopen.2021.47053

Lund LC, Hallas J, Nielsen H, et al. Post-acute effects of SARS-CoV-2 infection in individuals not requiring hospital admission: a Danish population-based cohort study. *The Lancet Infectious diseases* 2021;21(10):1373-82. doi: https://dx.doi.org/10.1016/S1473-3099(21)00211-5

Mainous AG, 3rd, Rooks BJ, Orlando FA. Risk of New Hospitalization Post-COVID-19 Infection for Non-COVID-19 Conditions. *Journal of the American Board of Family Medicine : JABFM* 2021;34(5):907-13. doi: https://dx.doi.org/10.3122/jabfm.2021.05.210170

McKeigue PM, McGurnaghan S, Blackbourn L, et al. Relation of incident Type 1 diabetes to recent COVID-19 infection: cohort study using e-health record linkage in Scotland. *medRxiv* 2022 doi: https://dx.doi.org/10.1101/2022.02.11.22270785

OpenSafely, Green A, Curtis H, et al. Describing the population experiencing COVID-19 vaccine breakthrough following second vaccination in England: a cohort study from OpenSAFELY. *BMC medicine* 2022;20(1):243. doi: https://dx.doi.org/10.1186/s12916-022-02422-0

Park C, Razjouyan J, Hanania NA, et al. Elevated Risk of Chronic Respiratory Conditions within 60 Days of COVID-19 Hospitalization in Veterans. *Healthcare (Basel, Switzerland)* 2022;10(2) doi: https://dx.doi.org/10.3390/healthcare10020300

Petersen EL, Gosling A, Adam G, et al. Multi-organ assessment in mainly non-hospitalized individuals after SARS-CoV-2 infection: The Hamburg City Health Study COVID programme. *European heart journal* 2022;43(11):1124-37. doi: https://dx.doi.org/10.1093/eurheartj/ehab914

Stastna D, Menkyova I, Drahota J, et al. To be or not to be vaccinated: The risk of MS or NMOSD relapse after COVID-19 vaccination and infection. *Multiple sclerosis and related disorders* 2022;65:104014. doi: https://dx.doi.org/10.1016/j.msard.2022.104014

##### Relevant studies carried out in non-OECD member countries

Abdelghani M, Atwa SA, Said A, et al. Cognitive after-effects and associated correlates among post-illness COVID-19 survivors: a cross-sectional study, Egypt. *The Egyptian journal of neurology, psychiatry and neurosurgery* 2022;58(1):77. doi: https://dx.doi.org/10.1186/s41983-022-00505-6

Agondi RC, Menechino N, Marinho AKBB, et al. Worsening of asthma control after COVID-19. *Frontiers in medicine* 2022;9:882665. doi: https://dx.doi.org/10.3389/fmed.2022.882665

Babtain F, Bajafar A, Nazmi O, et al. The disease course of multiple sclerosis before and during COVID-19 pandemic: A retrospective five-year study. *Multiple sclerosis and related disorders* 2022;65:103985. doi: https://dx.doi.org/10.1016/j.msard.2022.103985

Das S, Ray BK, Ghosh R, et al. Impact of COVID-19 pandemic in natural course of Moyamoya Angiopathy: an experience from tertiary-care-center in India. *Egyptian Journal of Neurology, Psychiatry and Neurosurgery* 2021;57(1):166. doi: https://dx.doi.org/10.1186/s41983-021-00412-2

Etemadifar M, Sedaghat N, Aghababaee A, et al. COVID-19 and the Risk of Relapse in Multiple Sclerosis Patients: A Fight with No Bystander Effect? *Multiple sclerosis and related disorders* 2021;51:102915. doi: https://dx.doi.org/10.1016/j.msard.2021.102915

Rahmani M, Moghadasi AN, Shahi S, et al. COVID-19 and its implications on the clinico-radiological course of multiple sclerosis: A case-control study. *Medicina clinica* 2022 doi: https://dx.doi.org/10.1016/j.medcli.2022.06.020

Soares FHC, Kubota GT, Fernandes AM, et al. Prevalence and characteristics of new-onset pain in COVID-19 survivours, a controlled study. *European journal of pain (London, England)* 2021;25(6):1342-54. doi: https://dx.doi.org/10.1002/ejp.1755

Xiong Q, Xu M, Li J, et al. Clinical sequelae of COVID-19 survivors in Wuhan, China: a single-centre longitudinal study. *Clinical Microbiology and Infection* 2021;27(1):89-95.

##### Excluded for not reporting severity/level of care received during acute phase of infection

Profili F, Seghieri G, Francesconi P. Effect of diabetes on short-term mortality and incidence of first hospitalizations for cardiovascular events after recovery from SARS-CoV-2 infection. *Diabetes Research and Clinical Practice* 2022;187:109872. doi: https://dx.doi.org/10.1016/j.diabres.2022.109872

Robineau O, Wiernik E, Lemogne C, et al. Persistent symptoms after the first wave of COVID-19 in relation to SARS-CoV-2 serology and experience of acute symptoms: A nested survey in a population-based cohort. *The Lancet regional health Europe* 2022;17:100363. doi: https://dx.doi.org/10.1016/j.lanepe.2022.100363

#### Appendix 4. Forest plots for new diagnoses of chronic conditions after SARS-CoV-2 infection, by age group and COVID-19 care type (inpatient vs. outpatient/mixed)

**Abbreviations:** CI = confidence interval; CVD = cardiovascular disease; IV = inverse variance; SE = standard error

##### Cardiovascular disorders

1. **Any cardiovascular disorder (CVD)**

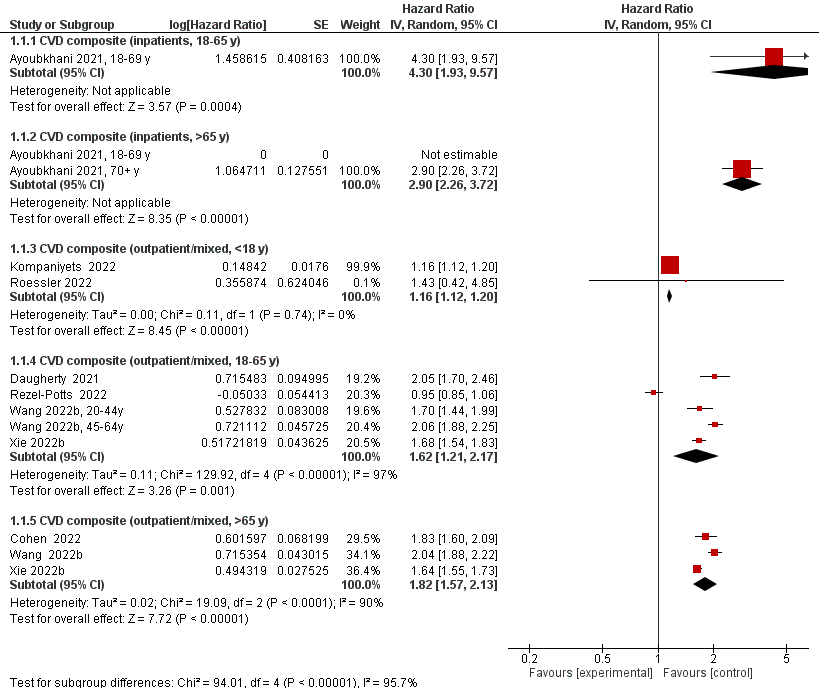

2. **Acute coronary disease (ACD)**

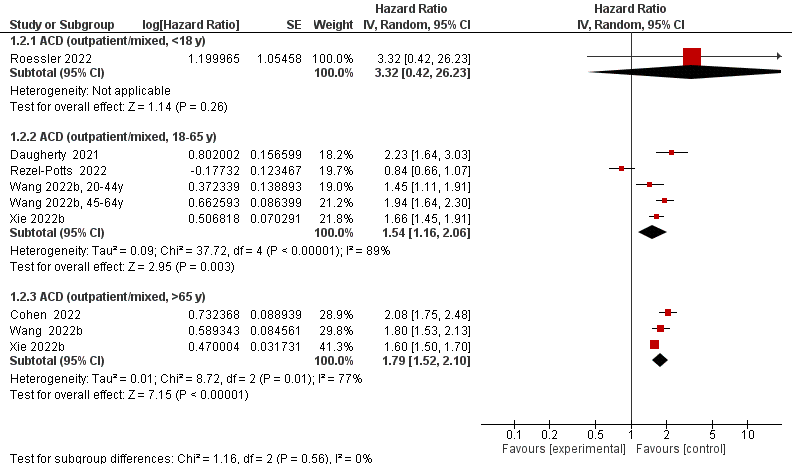

3. **Arrhythmias/dysrhythmias**

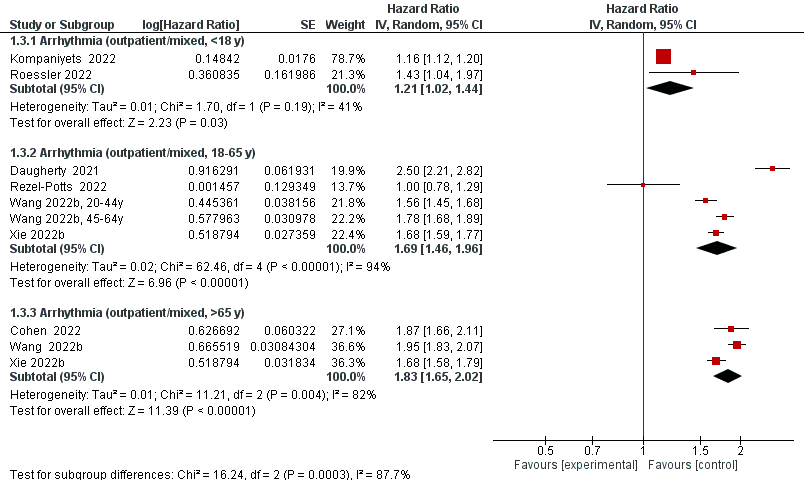

4. **Cardiomyopathy**

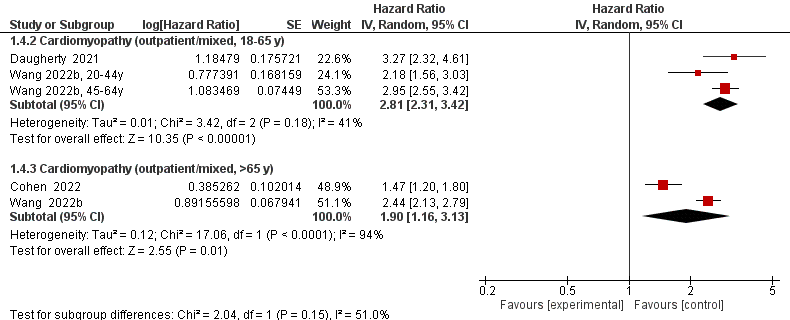

5. **Heart failure (HF)**

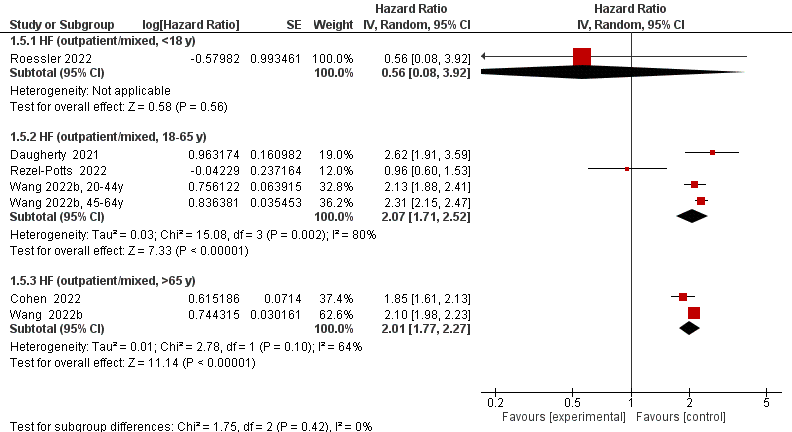

6. **Hypertension (HTN)**

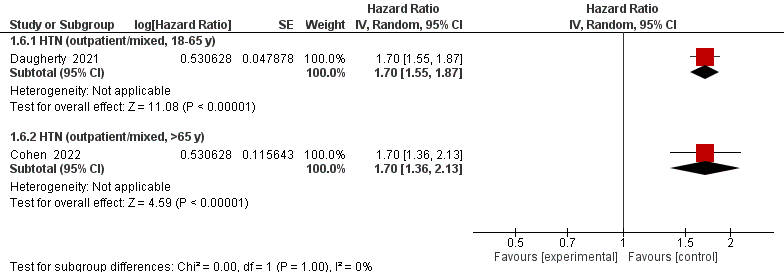

##### Chronic Kidney Disease

7. **Any chronic kidney disease (CKD)**

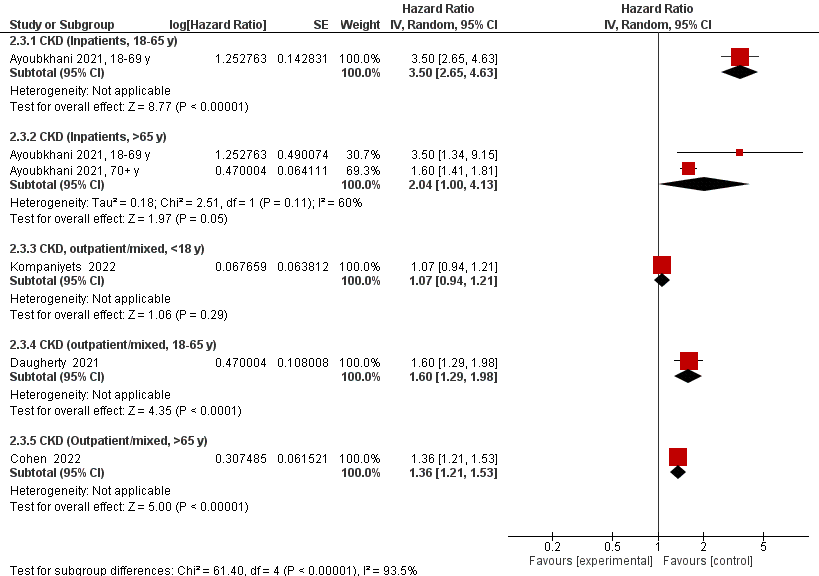

##### Diabetes

8. **Any diabetes**

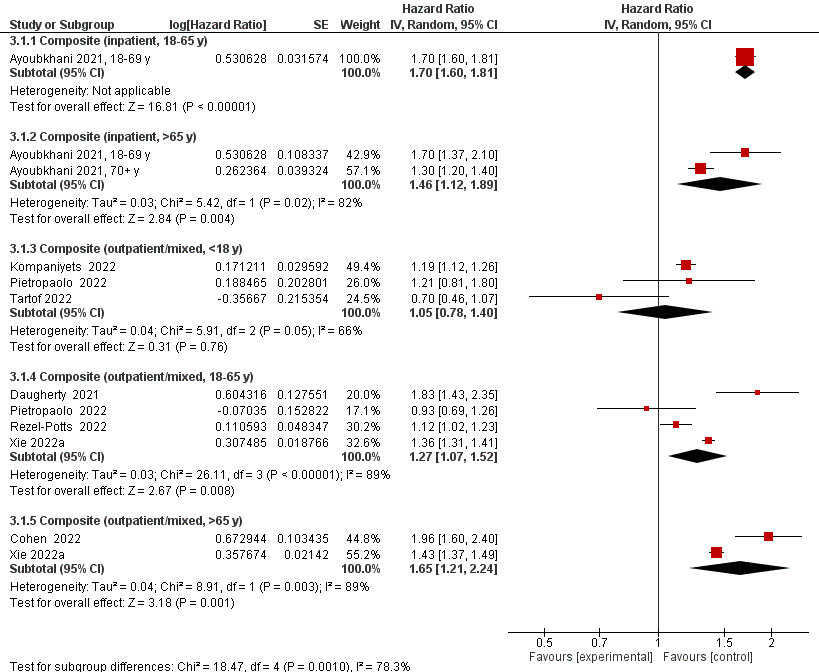

9. **Type 1 diabetes**

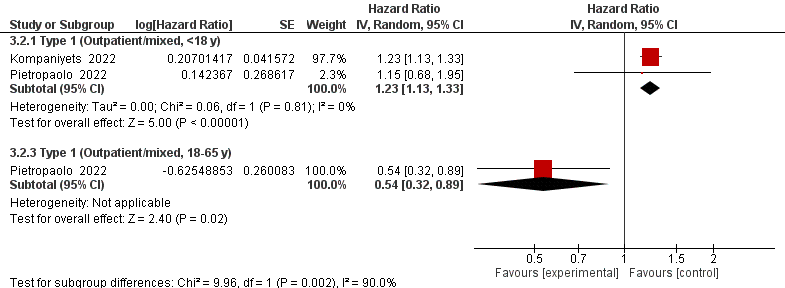

10. **Type 2 diabetes**

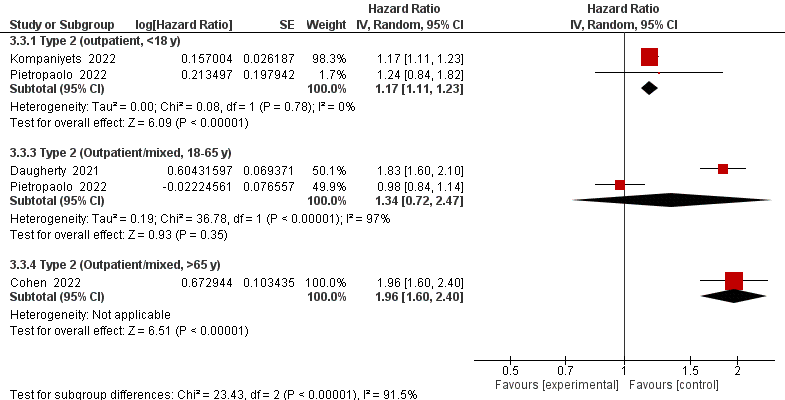

##### Mental Disorders

11. **Any mental disorder**

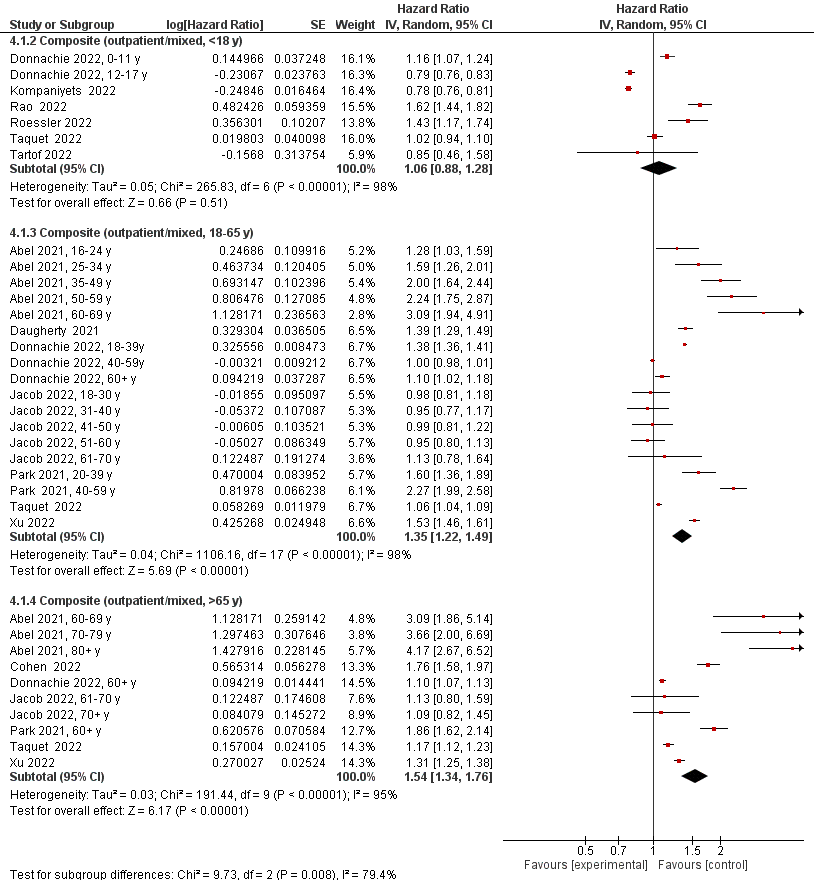

12. **Anxiety/anxiety disorders**

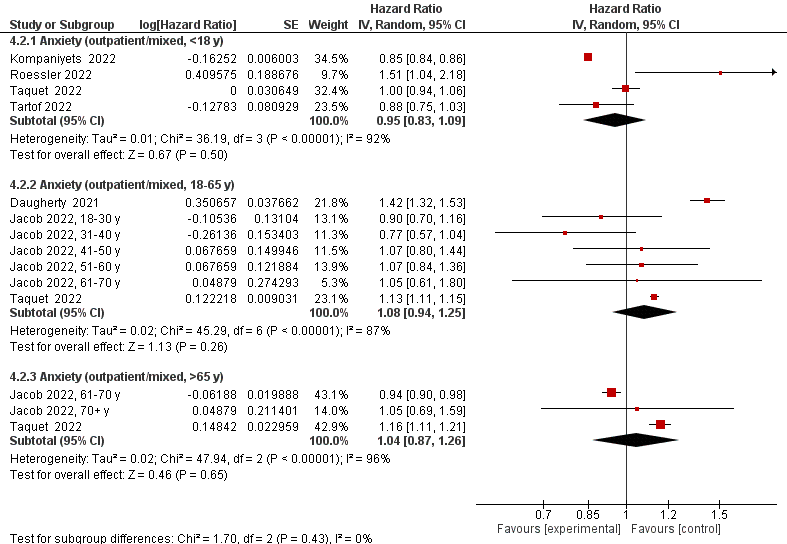

13. **Depression/depression disorders**

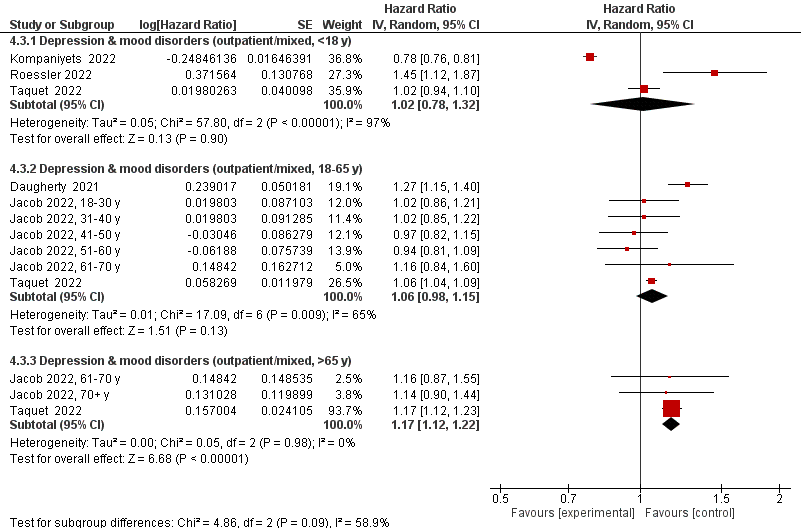

14. **Psychosis/psychotic disorders**

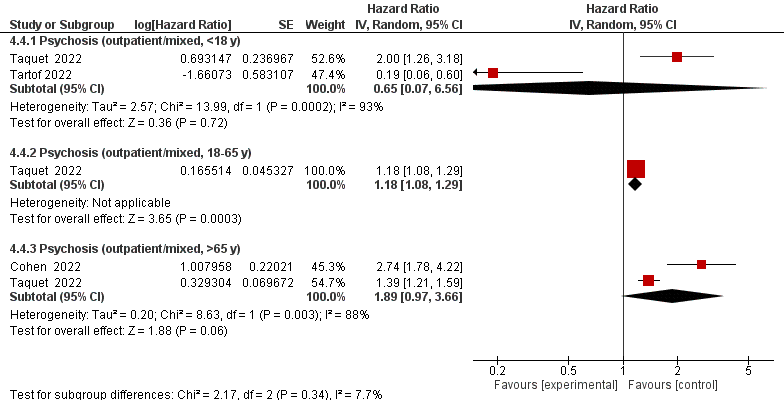

15. **Trauma and stress disorders**

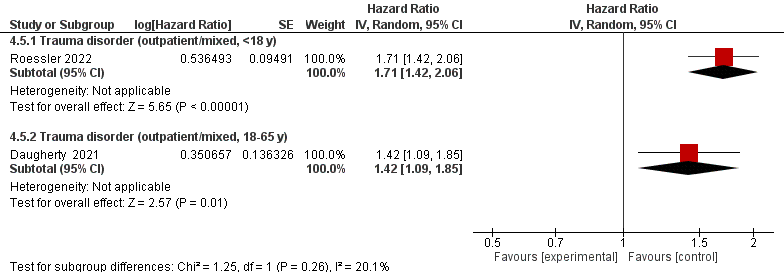

##### Musculoskeletal Disorders

16. **Any musculoskeletal (MSk) disorder**

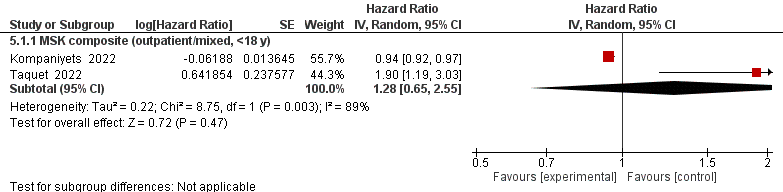

17. **Myoneural junction/muscle disease (MJD)**

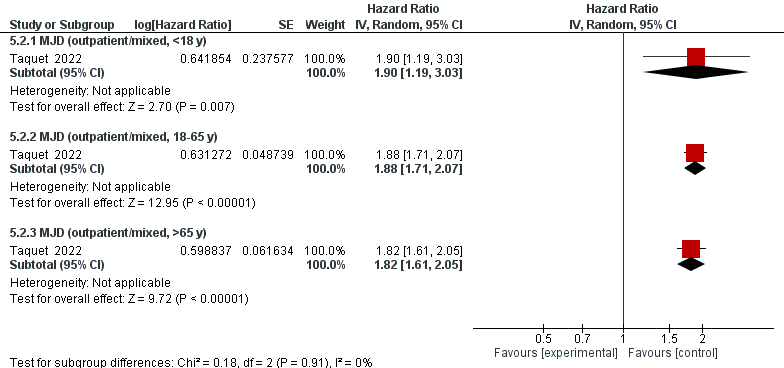

##### Neurological disorders

18. **Any neurological disorder**

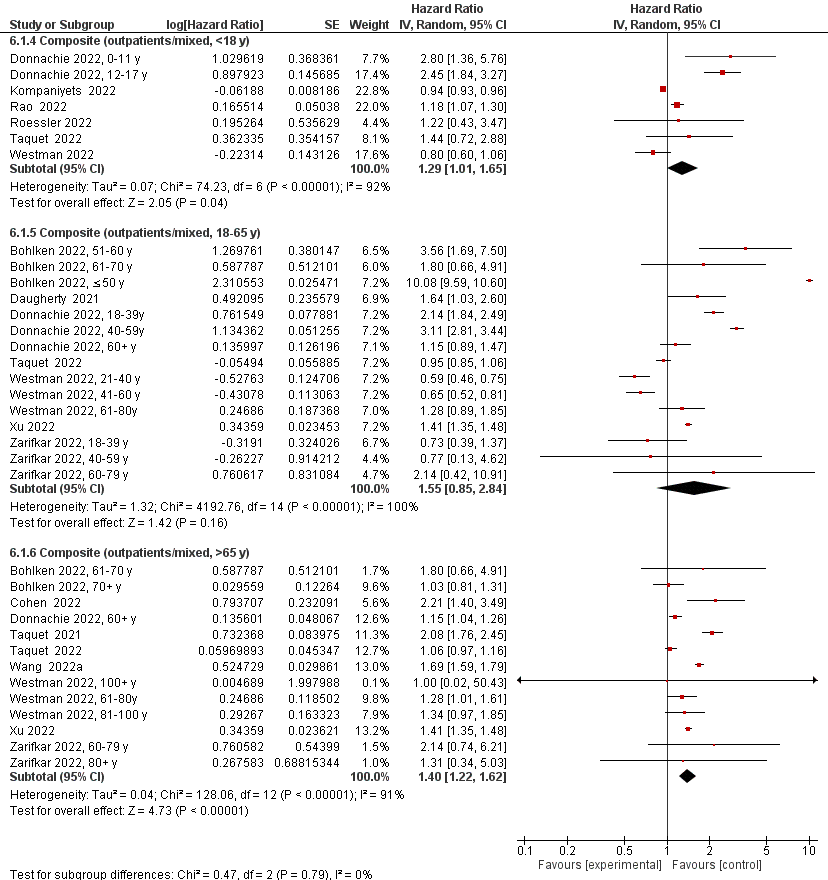

19. **Chronic fatigue syndrome (CFS)**

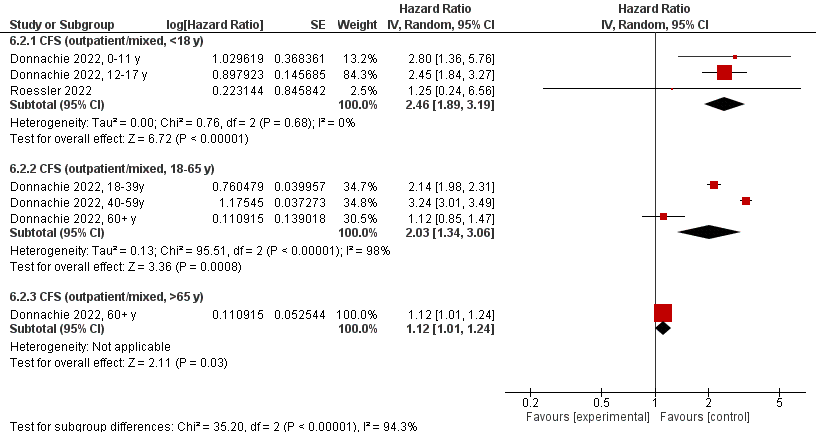

20. **Communication and motor disorders**

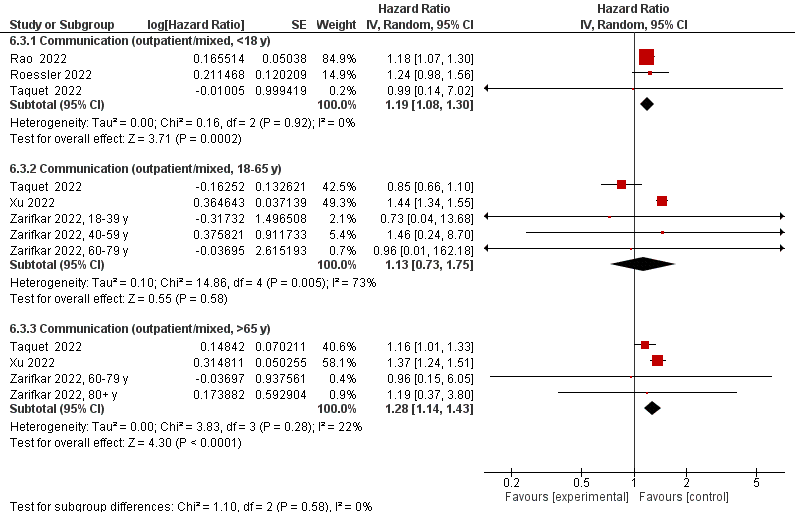

21. **Dementia/mild cognitive disorder**

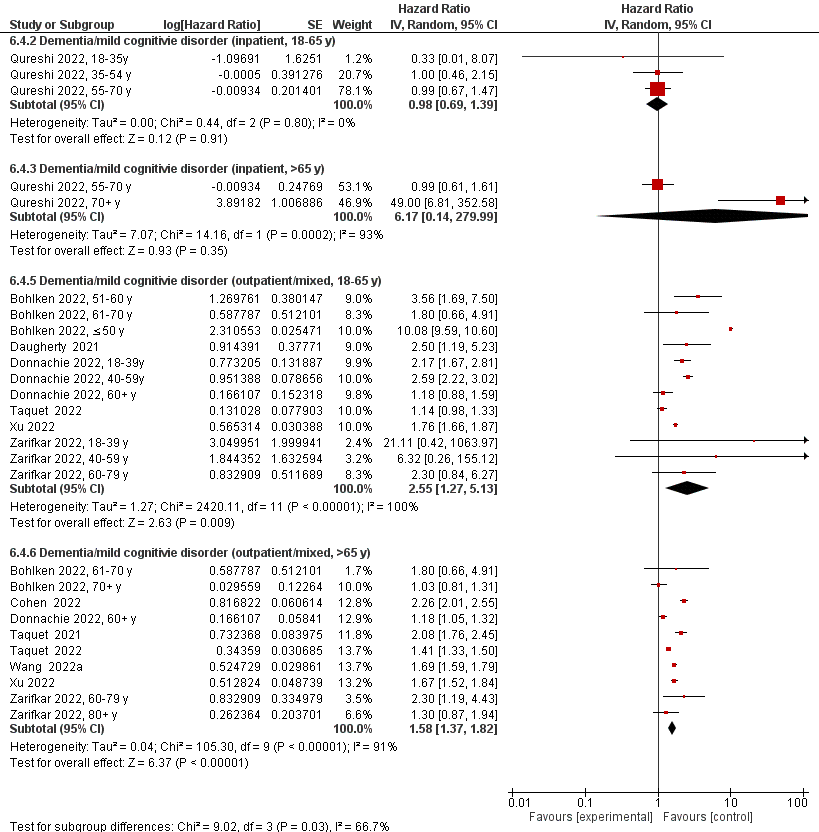

22. **Encephalopathy**

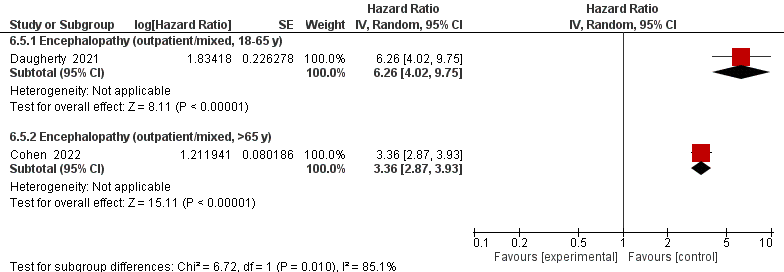

23. **Epilepsy**

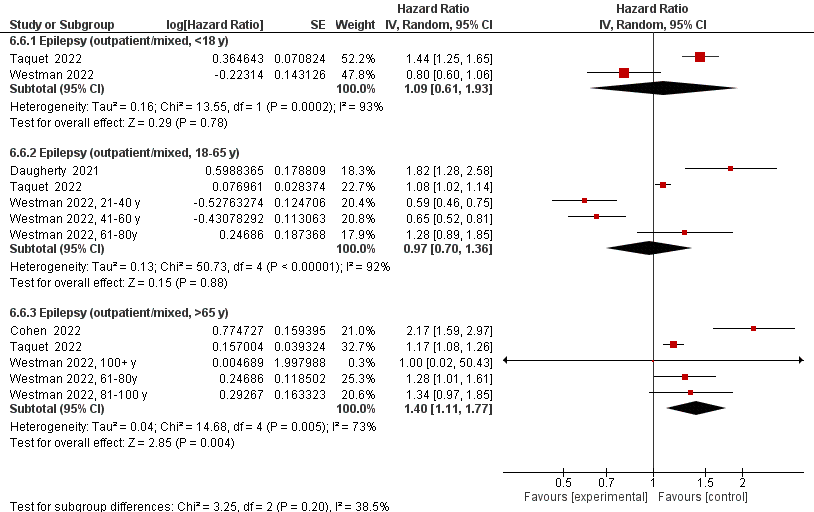

24. **Guillain-Barre syndrome (GBS)**

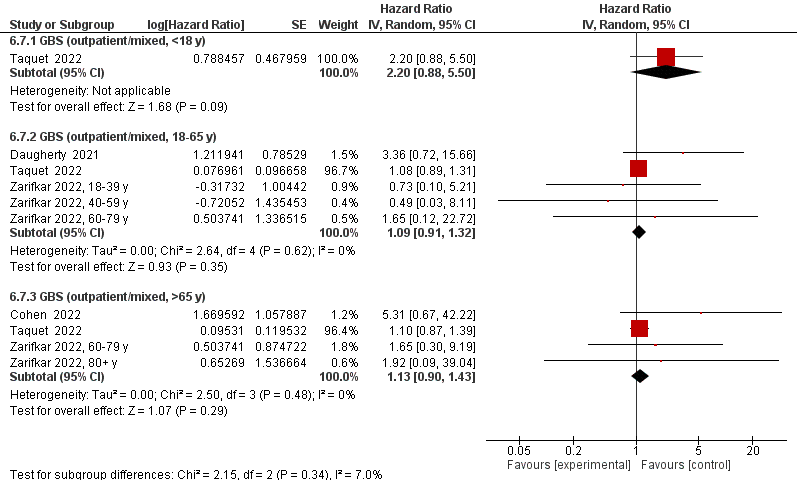

25. **Migraine**

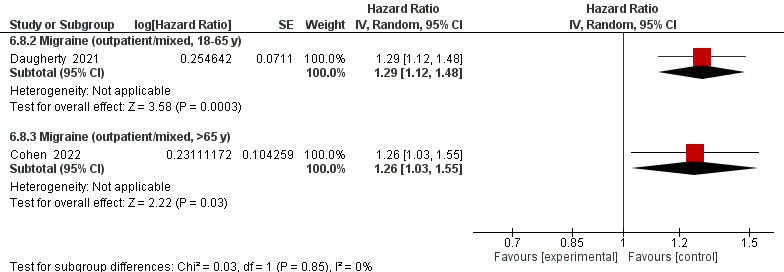

26. **Multiple sclerosis (MS)**

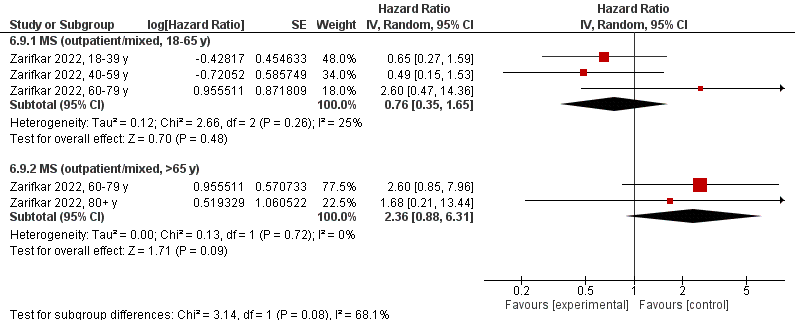

27. **Nerve disorders**

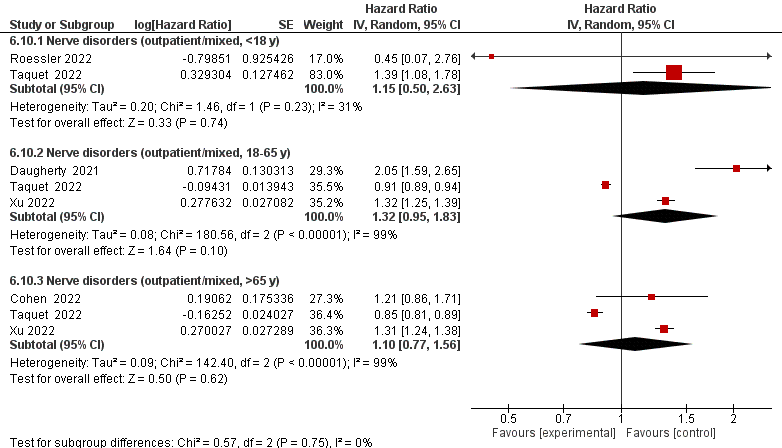

##### Respiratory Disorders

28. **Any respiratory (Resp) disorder**

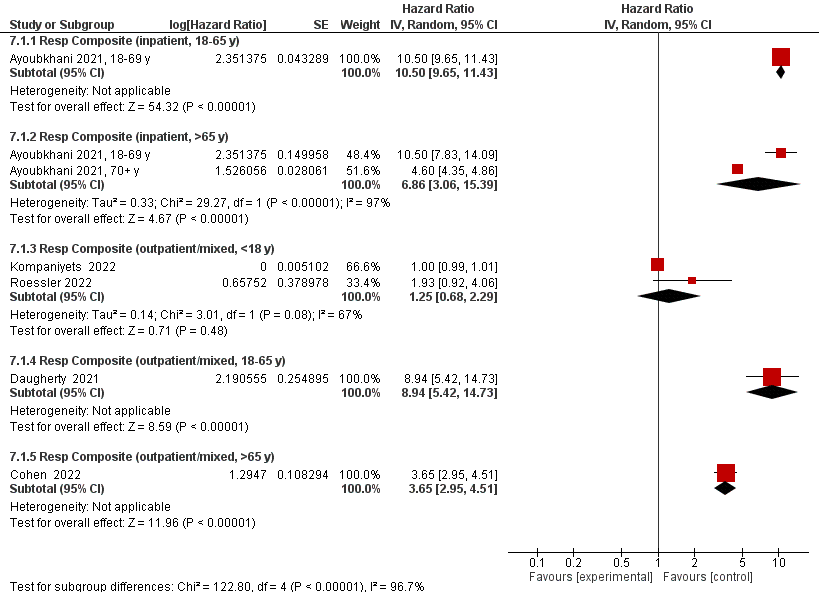

29. **Asthma**

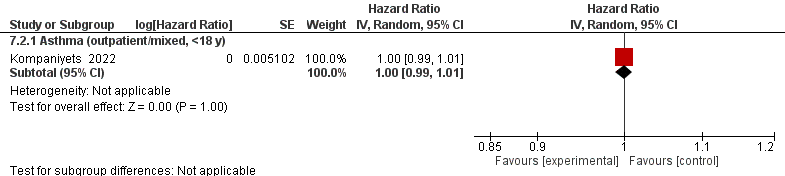

30. **Interstitial lung disease (ILD)**

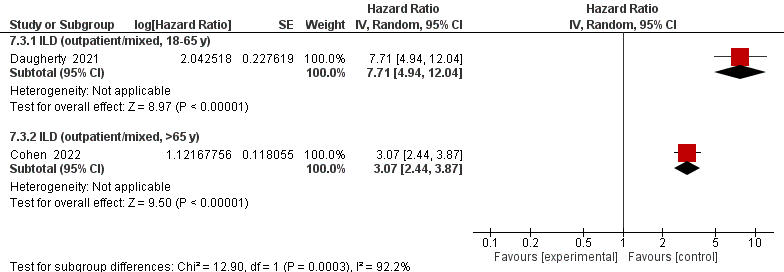

31. **Respiratory (resp) failure**

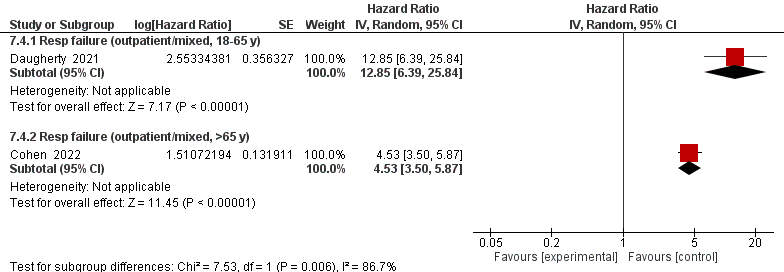

##### Stroke

32. **Any stroke**

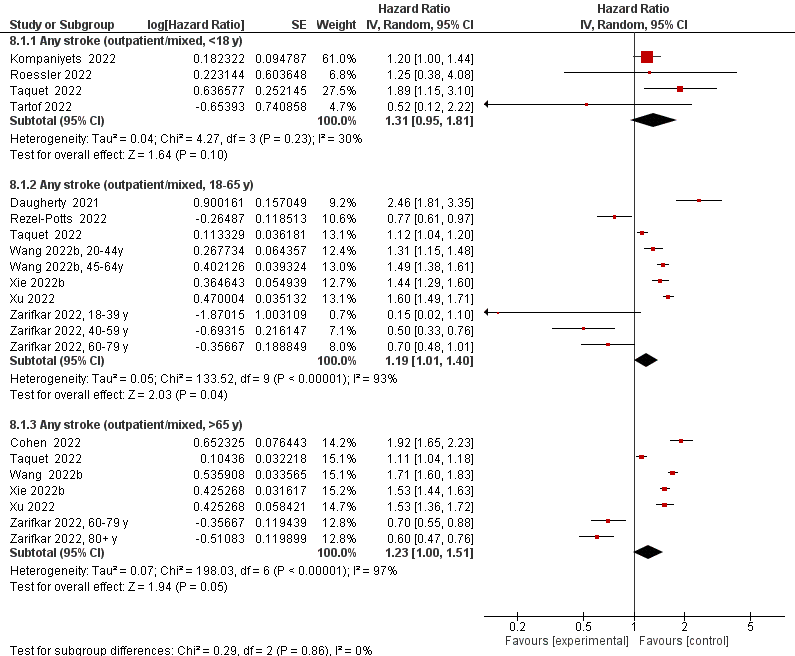

33. **Hemorrhagic stroke**

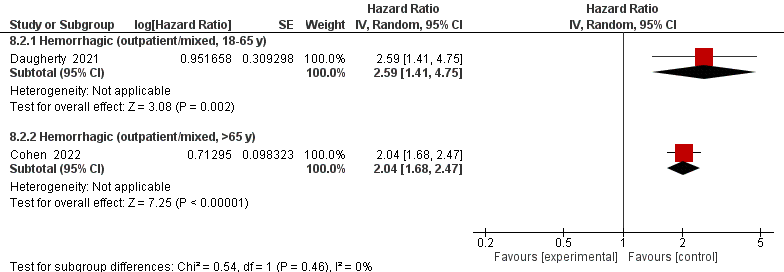

34. **Ischemic stroke**

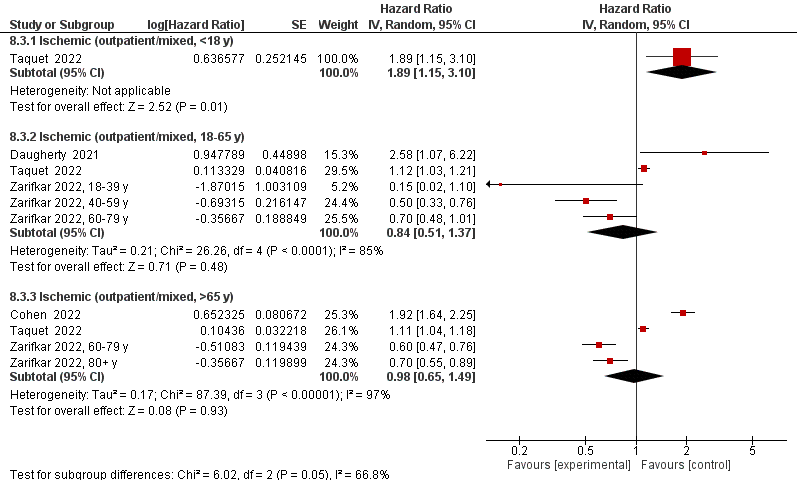

35. **Transient ischemic attack (TIA)**

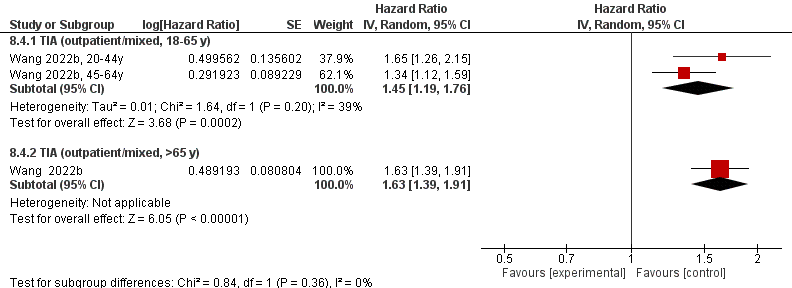
